# Individualized epidemic spreading models predict epilepsy surgery outcomes: a pseudo-prospective study

**DOI:** 10.1101/2023.03.16.23287370

**Authors:** Ana. P. Millán, Elisabeth C.W. van Straaten, Cornelis J. Stam, Ida A. Nissen, Sander Idema, Piet Van Mieghem, Arjan Hillebrand

## Abstract

Epilepsy surgery is the treatment of choice for drug-resistant epilepsy patients, but up to 50% of patients continue to have seizures one year after the resection. In order to aid presurgical planning and predict postsurgical outcome in a patient-by-patient basis, we developed a framework of individualized computational models that combine epidemic spreading with patient-specific connectivity and epileptogeneity maps: the Epidemic Spreading Seizure and Epilepsy Surgery framework (ESSES). The ESSES parameters were fitted in a retrospective study (*N* = 15) to reproduce invasive electroencephalography (iEEG)-recorded seizures. ESSES could not only reproduce the iEEG-recorded seizures, but significantly better so for patients with good (seizure-free, SF) than bad (non-seizure-free, NSF) outcome (area under the curve *AUC* = 0.73). Once the model parameters were set in the retrospective study, ESSES can be applied also to patients without iEEG data. We illustrate here the clinical applicability of ESSES with a *pseudo-prospective study* (*N* = 34) with a blind setting (to the resection strategy and surgical outcome) that emulated the presurgical conditions. ESSES could predict the chances of good outcome after *any* resection by finding patient-specific optimal resection strategies, which we found to be smaller for SF than NSF patients, suggesting an intrinsic difference in the network organization or presurgical evaluation results of NSF patients. The actual surgical plan also overlapped more with the optimal resection, and had a larger effect in decreasing modeled seizure propagation, for SF patients than for NSF patients. Overall, ESSES could correctly predict 75% of NSF and 80.8% of SF cases pseudo-prospectively. Our results show that individualised computational models may inform surgical planning by suggesting optimal resections and providing information on the likelihood of a good outcome after a proposed resection. This is the first time that such a model is validated on a fully independent cohort without the need for iEEG recordings.

## 1 Introduction

Surgical resection is often the most effective treatment to achieve seizure control for patients with drug-resistant focal epilepsy. The surgery requires the generation of an hypothesis of the epileptogenic zone (EZ) by means of extensive presurgical evaluations, and its subsequent removal or disconnection during surgery [1]. Despite extensive investigations, there has only been a slight improvement in prognosis over the past two decades [2, 3], and between 30 to 50% of the patients who undergo surgery continue to have seizures one year later, depending on etiology and location of the EZ [4]. A key conceptual change in recent years is the notion of *epileptogenic networks*, which takes into account the complex interplay between different brain regions in promoting and inhibiting seizure generation and propagation [5–7]. As a consequence, the effect of a given surgery is to be measured against the whole epileptogenic network: a small resection involving heavily connected regions may have widespread effects, but it may also be compensated for by the remaining network [8, 9]. Several network-based studies have found group-level differences between seizure-free and non-seizure-free patients [9–11], with removal of pathological hub (i.e. central) regions typically associated with seizure-freedom [12]. These results highlight the need to consider *patient-specific connectivity* [13] in order to tailor the surgery to each patient [14].

A data-driven manner to study the relation between individual brain networks and surgical outcomes involves *computational models of seizure dynamics*, which allow us to simulate seizure propagation *in silico*. Different resection strategies can be tested on the computational model before the actual surgery [15–24]. The models can be fitted to patient-specific data of the brain structure and seizure dynamics, allowing us to tailor the resection strategy for each patient. Within this perspective, previous studies have obtained remarkable success at a group level: Sinha et al. [22] found that the removal of regions identified as epileptogenic according to an EEG-brain network dynamical model predicted surgery outcome with 81.3% accuracy. Proix et al. [25] using a seizure model known as the *epileptor* [26] based on MRI (magnetic resonance imaging) connectivity, found significant differences (*p <* 0.05) between patients with good (Engel class I) and bad (Engel class III) outcomes at the group level. Subsequent studies also found a better match between the hypothesized epileptogenic and propagation zones for seizure-free than non-seizure-free patients [27, 28]. Similarly, Sip et al. [29] simulated patient-specific resection strategies by means of a *virtual resection*, and found that virtual resections in their model correlated with surgical outcome, with larger effects found for patients with good outcome (Engel Classes I and II). In an independent study, Goodfellow et al. [19] also found significant differences in the model prediction between Engel Class I and class IV patients, using an electrocorticogram modeling framework.

Following the same rationale, we developed a computational model of seizure propagation and epilepsy surgery based on epidemic spreading dynamics and patient-specific MEG brain connectivity [30], to which we refer here as the *Epidemic Spreading Seizure and Epilepsy Surgery model (ESSES)*. Epidemic models describe the spread of an infectious agent through a network. Epidemic processes on fixed networks have a rich mathematical history [31] with a plethora of models that can be exploited for epilepsy surgery optimization [24, 30]. Although such models ignore the underlying bio-physical processes that lead to seizure generation and propagation, they do describe the basic rules that govern spreading processes. In previous studies [30, 32], we found that epidemic spreading models could reproduce stereotypical patterns of seizure propagation as recorded via invasive electroencephalography (iEEG) recordings. Moreover, once fitted with patient-specific data, ESSES could identify alternative resection strategies, either of smaller size or at a different location than the actual surgery [24, 30]. In a more recent study [32], we showed that the goodness-of-fit of the ESSES seizures and those recorded via iEEG predicted surgery outcome –with an area under the curve of 88.6% – indicating that ESSES not only reproduces the basic aspects of seizure propagation, but it also captures the differences, either in the location of the resection area relative to the EZ, or intrinsically in the iEEG or MEG data, between patients with good and bad outcome. Importantly, the global parameters of ESSES were defined at the population level, and the model was individualized for each patient via patient-specific MEG networks, which characterized the local spreading probabilities. As a consequence, ESSES can be extended to patients without iEEG recordings, in contrast to previous modeling studies, which typically required the existence of patient-specific iEEG data to individualize the model for each patient [22, 25, 27, 33–36]. IEEG allows for a highly resolved description of seizure dynamics, but its spatial sampling is sparse and it is highly invasive. Consequently, it is only part of the presurgical evaluation in a selection or patients.

Here we performed a pseudo-prospective blind study (34-patient validation cohort) to validate the clinical applicability of ESSES to a) identify optimal resection strategies and b) predict the likelihood of a good outcome after a proposed resection strategy, in a patient-by-patient basis. In order to emulate the clinical presurgical conditions, the research team was blind to the postsurgical data of the patients, namely the resection area and surgical outcome, during the ESSES analyses, and the multimodal presurgical information available for each patient was integrated into ESSES. ESSES can identify optimal resection strategies, as well as simulate the effect of a given resection *in silico*. Within this set-up, we tested three hypotheses: a) SF patients would have smaller optimal resections, b) SF patients would have a larger overlap between optimal and planned (clinical) resections, and c) the planned resection would have a larger effect (in ESSES) for SF than NSF patients. We found that these three ESSES biomarkers, namely the size of the optimal resection, their overlap with the planned resection, and the effect of the planned resection on ESSES seizures, provided estimates of the likelihood of a good outcome after the surgery, as well as suggesting optimal (alternative) resection strategies. We envisage that the implementation of a modeling scheme such as ESSES in clinical practice may inform the planning of epilepsy surgery. Different surgical plans can be tested in the ESSES model for each patient, such that strategies that lead to a large decrease of propagation in the model are more likely to lead to seizure freedom. ESSES may also find optimal (alternative) resection strategies, in cases where ESSES predicts a bad outcome with the planned resection. Optimal strategies can then lead to new surgical plans, whose effect can then be tested in ESSES again.

## 2 Results

Here we validate the clinical applicability of ESSES to **A)** identify optimal resection strategies that may improve surgical outcomes and **B)** provide estimates of the probability of postsurgical seizure freedom, given a surgical plan. The key goal of ESSES is to identify surgical candidates who would have a bad outcome (NSF patients) so that the surgical plan can be adjusted. This study combined a retrospective analysis on a *modeling cohort* (*N* = 15) that was used to set the model hyperparameters, and a pseudo-prospective study on a *Validation cohort* (*N* = 34) to validate the ESSES findings and to emulate its clinical application in a blind set-up that mimics the clinical presurgical conditions. The researchers were blind to the performed surgery and surgical outcome during the application of ESSES to the validation cohort.

The study was performed as follows:

1. **Seizure model: definition and fitting (modeling cohort)**. An SIR-type of epidemic spreading process models seizure propagation over patient-specific brain connectivity. iEEG data from the modeling cohort was used to fit the global parameters of the spreading model so that the ESSES seizures matched those recorded via iEEG, as shown in figure 1A.
2. **Individualized ESSES framework: patient-specific models**. ESSES was individualized for each patient: patient-specific MEG brain connectivity defines the network on which ESSES computes seizure propagation. Multi-modal patient-specific data, available from presurgical evaluations,defined the seed regions (i.e. the seizure onset regions) based on *epileptogenicity* or *seed-probability maps*.
3. **Alternative resection strategies (aim A)**. ESSES incorporates an optimization algorithm to determine the optimal resection strategies for each patient. These acted as a benchmark against which the planned resection for each patient could be tested.
4. **Simulation of the planned resection strategy (aim B)**. The resection plan of each patient was simulated in ESSES with a virtual resection that emulated the actual surgical, and the subsequent decrease is seizure propagation was measured.
5. **Statistical analyses (aim B)**. We compared the ESSES predictions (steps 3 and 4) between patients with good and bad outcome. We defined the NSF class as the positive class for classification and prediction testing.

**Figure 1:**
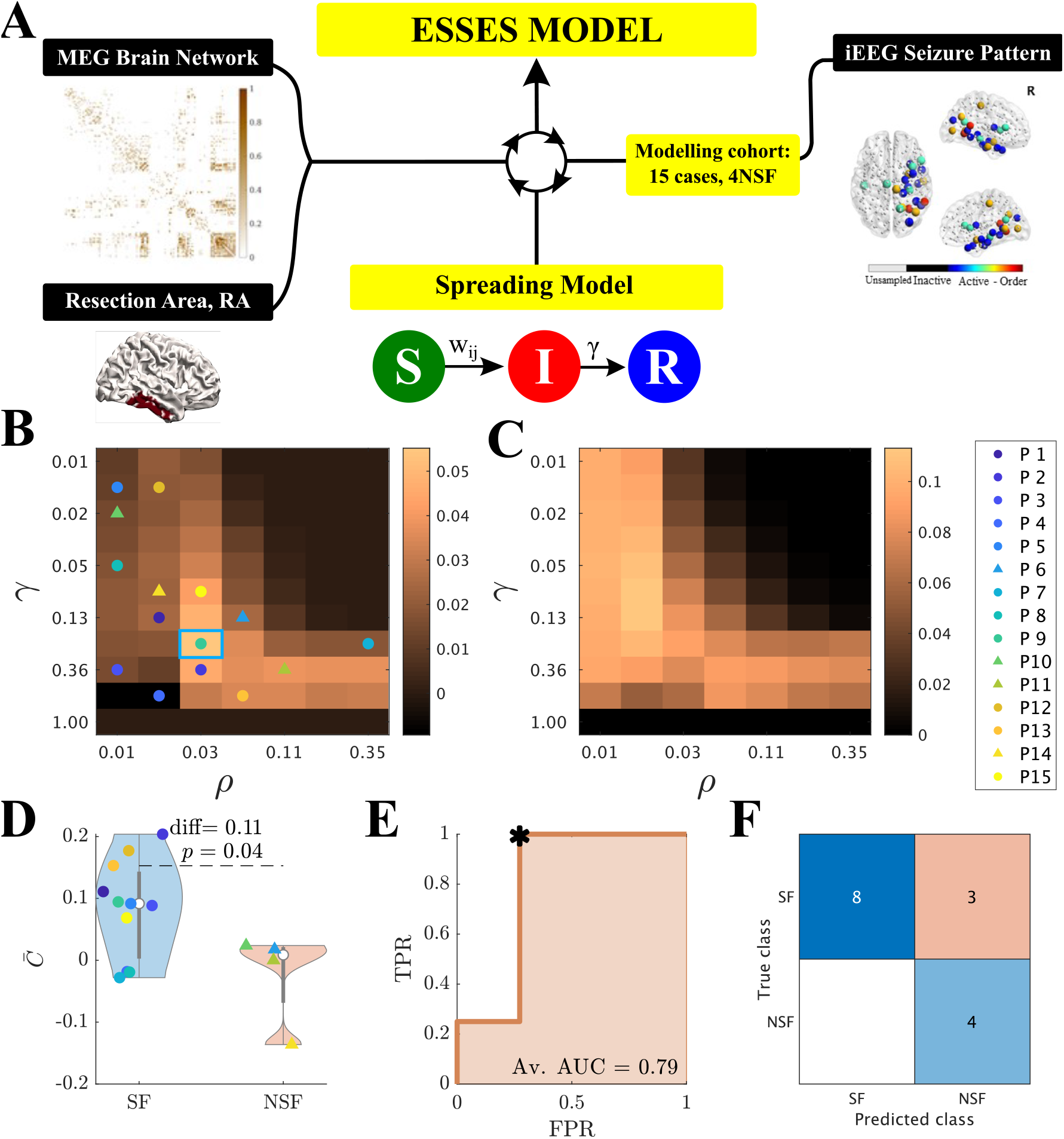
**A** Sketch of the ESSES parameter-fitting scheme. The parameters controlling seizure propagation in ESSES, namely the density of links in the network *ρ* and the global recovery probability *γ* were set so as to maximize the similarity between the ESSES seizures and those recorded via iEEG (eq. 1). For the patients of the modeling cohort, seizures were simulated using the SIR dynamics over the MEG patient-specific brain network and using the resection area as the seed of epidemic spreading. The resulting seizures were then compared to those recorded via iEEG. **B, C** 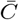 (*ρ, γ*) maps displaying the average model fit (**B**) and the corresponding standard deviation (**C**) for the modeling cohort. The data points in the left panel indicate the parameters corresponding to the best individual fit for each patient, with circles (triangles) indicating SF (NSF) cases (corresponding *C* values can be seen in Supp. figure 2). The blue square marks the maximum of the goodness-of-fit, and the corresponding (*ρ, γ*) values were used for the subsequent analyses. The y-axis is shown using a logarithmic scale. **D** Violin plots of the goodness-of-fit distributions for the SF and NSF groups. **E** ROC (receiving-operator-characteristic) curve analysis, where TPR and FPR respectively indicate the true positive (NSF patient classified as NSF) and false positive (SF patient classified as NSF) rates. The optimal classification point (Youden criterion) is shown by a black asterisk, and the corresponding confusion matrix is shown in panel **F**. The confusion matrix indicates the number of SF and NSF cases that were correctly (diagonal elements) and incorrectly (off-diagonal elements) classified.

This analysis pipeline was first implemented in the modeling cohort on a retrospective study that served to set all model hyperparameters. Then, steps 2 *−* 5 were applied to the validation cohort in a pseudo-prospective study with a blind set-up. The modeling and analysis schemes are outlined in figure 2.

**Figure 2:**
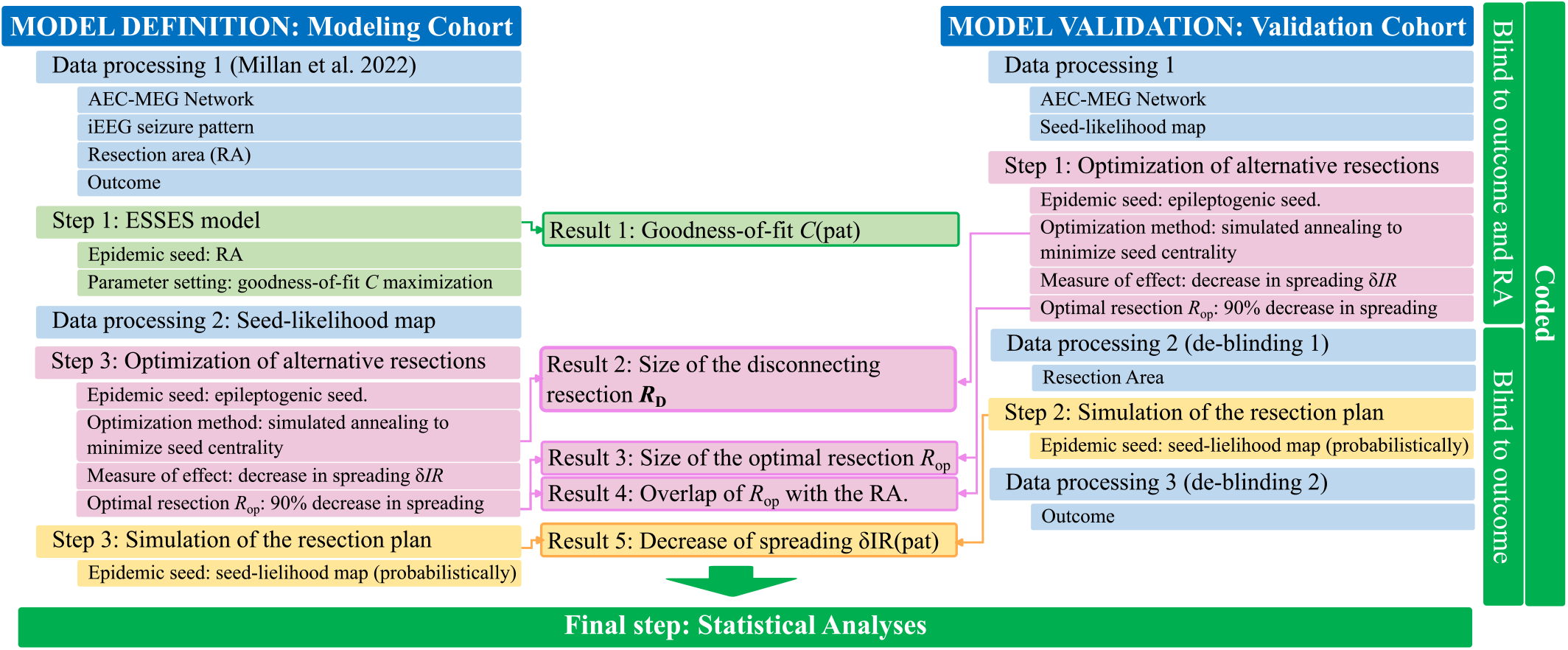
Processing and analysis pipeline.

### 2.1 Seizure propagation as an epidemic spreading process

We modeled seizure propagation by a Susceptible-Infected-Recovered (SIR) epidemic process, where the S-I-R states account respectively for the healthy (pre-ictal), ictal and healthy (post-ictal) states, coupled with patient-specific brain connectivity (derived from MEG data) to define the local spreading probabilities. The SIR model describes the spread of an infection from an initial set of infected nodes, the seed regions, to the other nodes in the network, and the recovery of the infected nodes, without re-infections [31, 37]. Here we confined to one of the simplest compartmental SIR models, using a discrete-time setting where the spreading probability from node *i* to node *j* corresponded to the coupling strength *w*_*ij*_ on the patient-specific brain network and where the recovery probability *γ* was set to be equal for all nodes. The brain network was initially thresholded (by setting the weakest links to zero) at different densities *ρ* indicating the fraction of non-zero links remaining in the network after thresholding (see Methods sections 5.2 and 5.5, and Supp. section 5).

The two control parameters of the ESSES model are thus the global recovery probability *γ* and the network density *ρ*. Using the iEEG data for the modeling cohort patients, we fit *γ* and *ρ* to reproduce the iEEG-recorded seizure propagation patterns with the ESSES-modeled seizures, setting the resection area was used as the epidemic seed (see 5.5 and Supp. section 5.2 for more details) [30, 32]. The degree of similarity between the ESSES and iEEG seizures was measured with the *goodness-of-fit C*(*ρ, γ*) as defined in eq. 1. The resulting diagram resembled a familiar phase transition (figure 1B), with an interface of high goodness-of-fit (yellow regions) corresponding to a roughly constant spreading-to-recovery ratio *ρ/γ* = const, in agreement with other studies [38]. Most individual best fits (data-points in figure 1B) fell within this region, although there was large variability among the individual fit maps (in fact, we found low signal to noise ratios of approx. 1*/*2, see figure 1C).

The maximum goodness-of-fit is indicated by a blue square in figure 1B, and sets the working point of ESSES for the remaining analyses. At this working point, the SF group presented a significantly better fit than the NSF group (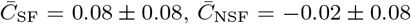, *C*_SF_ *− C*_NSF_ = 0.11, rank-sum *rks* = 72, *p* = 0.04), as shown in figure 1D. A ROC classification analysis indicated a good classification (AUC = 0.79) between the SF and NSF groups based (figure 1E). At the optimal classification point (Youden criterion, figure 1F), all NSF patients were correctly identified in this case (accuracy = 0.80, precision = 0.57, sensitivity = 1.00 and *F* 1 = 0.71). The high sensitivity suggests that all patients identified as SF by ESSES could proceed to surgery with high expectations (100% in this group) of a good outcome. On the contrary, patients identified as NSF should be examined further (e.g. by performing further presurgical evaluations or considering other resection plans) as they had a 57% chance of bad outcome with the proposed surgery (to be compared with a 26% chance of bad outcome expected simply from the relative group sizes).

### 2.2 Presurgical hypothesis of the seed regions

A key ingredient of ESSES is the definition of the epileptogenic or seed regions. Here we defined epileptogenicity or seed-propbability maps *SP*_*i*_ indicating the probability that each brain region *i* gave rise to a seizure The *seed-probability* maps integrated the patient-specific multimodal presurgical information (encoded in the local patient database [39]) in a quantitative and systematic manner, that was adapted for each patient to include the data from the presurgical evaluations that they had undergone (see Methods section 5.6 and Supp. section 4 for details). The resulting seed-probability maps for two representative patients from the modeling cohort are illustrated in figure 3B,D together with the corresponding resection areas (panels A, C). The seed-probability maps show wider spatial patterns than the resection areas, and may involve several lobes in both hemispheres. The resection areas in the two cases shown here were contained within the most likely seed regions. In general, the resection areas had a larger seed-probability than expected by chance for all patients. We did not find significant differences in the overlap between the resection areas and the seed-probability map between SF and NSF patients (see Sup. Fig. S1 and S2).

**Figure 3:**
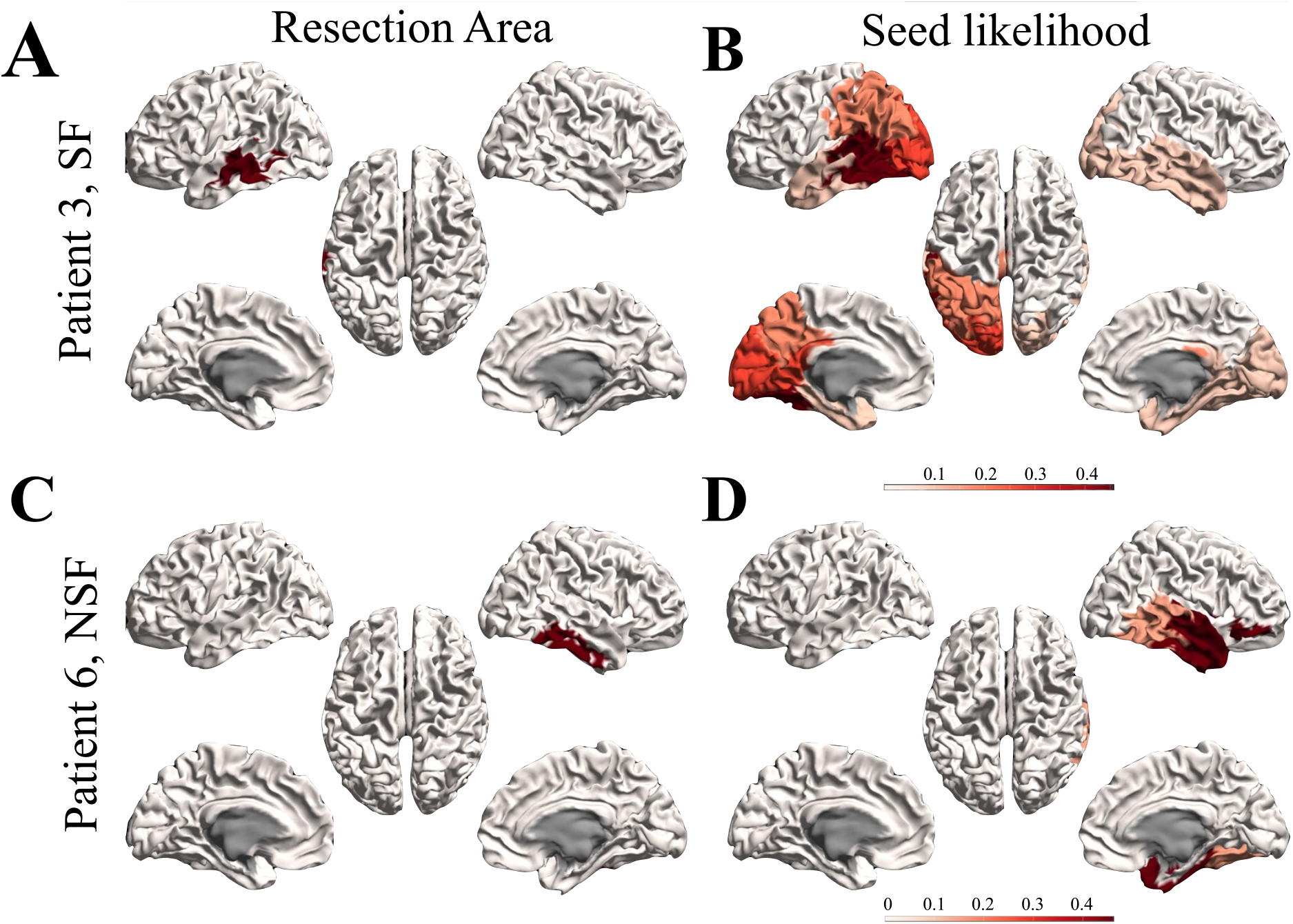
Seed-probability maps. Resection areas (left) and seed-probability maps (right) as derived from the database with presurgical information for two representative cases from the modeling cohort: patient 3 (SF, top) and 6 (NSF, bottom).

### 2.3 Optimal resection strategies

ESSES can derive individualized optimal (alternative) resection strategies via an optimization algorithm based on simulated annealing. The optimization algorithm was first designed using the modeling cohort data (see Methods section 5.7), and was then applied to the validation cohort in a blind setting. Here we report on the results for the validation cohort, corresponding analyses for the modeling cohort can be found in the supplementary information (Supp. section 5.3 and Supp. figures 3 and 4).

The optimization algorithm searched for resections *R* of increasing size *S*(*R*) that minimized the *seed efficiency E*_*R*_(seed), exploiting the link between epidemic spreading dynamics and network structure. To diminish differences due to seed extent and initial efficiency, we defined the normalized seed efficiency *e*_*R*_(seed) = *E*_*R*_(seed) *− E*_0_(seed), where *E*_0_(seed) is the initial seed efficiency (figure 4A). *e*_*R*_(seed) decreased with the size of the resection for all patients. At the group level, the SF group showed a significantly larger *e*_*R*_(seed) than the NSF group (repeated measures ANOVA test, *F* (19) = 37.95, *p <* 10^*−*89^), for all considered seed sizes except *S*(*R*) = 1. Moreover, the effect of increasing the resection size on *e*_*R*_(seed) was larger for the SF than the NSF group (*F* (19) = 3.78, *p <* 10^*−*6^).

**Figure 4:**
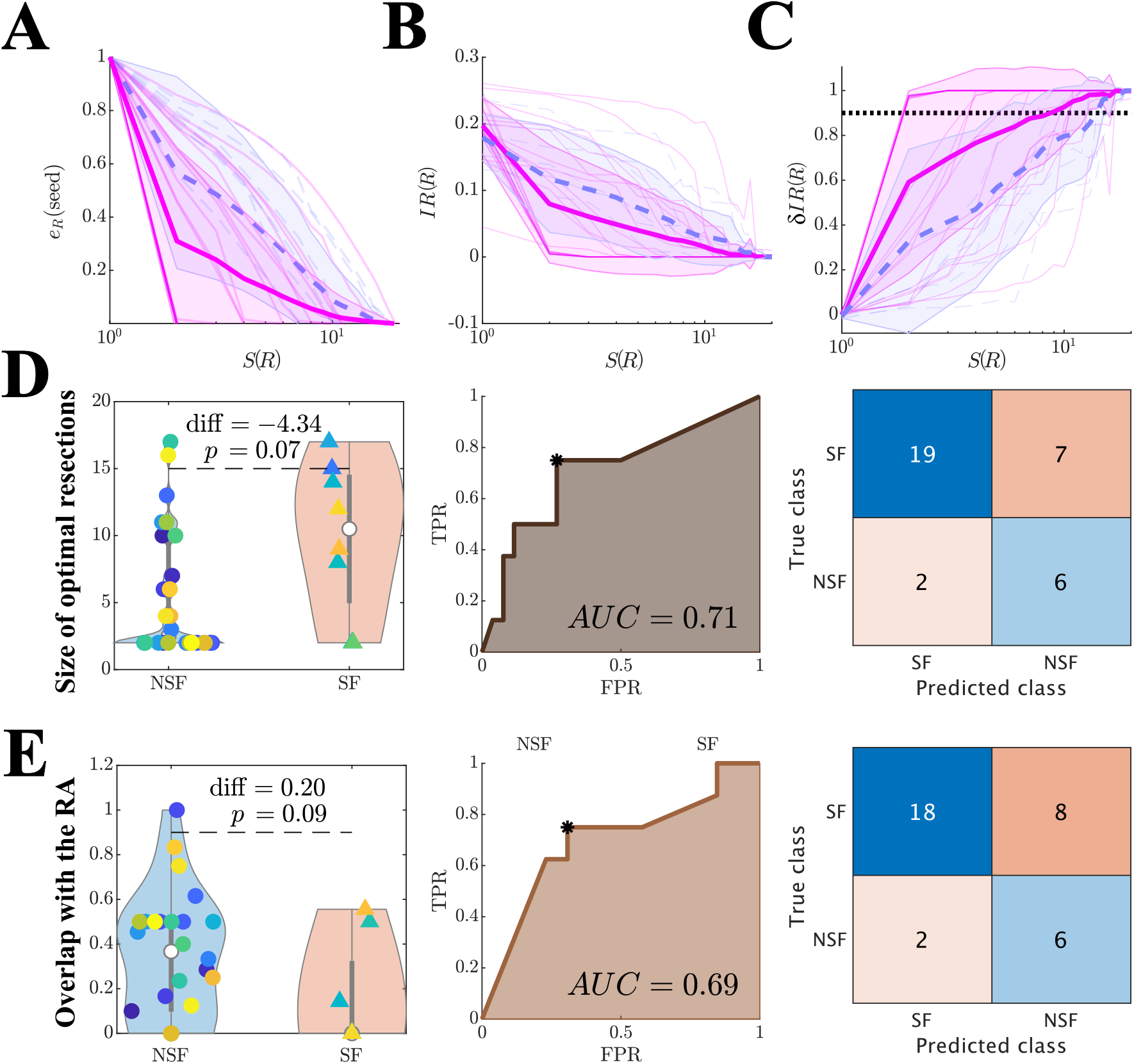
Optimal (alternative) resection strategies (validation cohort). Effect of optimal virtual resections of size *S*(*R*) on **A** normalized seed efficiency *e*_*R*_(seed), **B** seizure propagation after the resection *IR*(*R*), and **C** normalized decrease in seizure propagation *δIR*(*R*). Blue dashed lines stand for NSF patients, and pink solid lines for SF patients. Thin lines show individual patients, and darker wide lines the group averages, with shaded areas indicating the standard deviations. **D**,**E** Group level comparison of the size of optimal resections *S*(*R*_*op*_) (**D**) and their overlap with the resection area *Ov*(*R*_*op*_, *R*_*D*_) (**E**). The left panels show the distribution of values of each patient group, with significance results obtained with exact two-sided Wilcoxon ranksum tests. The center panels show the corresponding ROC classification analyses, where TPR and FPR stand respectively for the true positive and false positive rates. Finally, the right panels show the confusion matrices corresponding to the optimal point (Youden criterion, black asterisks in the middle panels) of the ROC curves.

The expected effect of a resection *R* on seizure propagation was quantified by comparing seizure propagation *IR* before (*IR*(0)) and after (*IR*(*R*)) the resection. Seizure propagation depended heavily on the seed realization such that a bi-stable regime emerged where, depending on the seed realization, the ESSES seizures either propagated macroscopically or died locally (an exemplary case is shown in Supp. figure 3). Thus, results reported here were averaged over 300 independent realizations of the seed regions and SIR dynamics. The SF group presented a significantly larger decrease in seizure propagation (*F* (19) = 16.65, *p <* 10^*−*43^), than the NSF group. In this case the effect of increasing the resection size was not significantly different between the two groups (*F* (19) = 1.43, *p* = 0.11).

We defined the *normalized decrease in seizure propagation δIR*(*R*) = (*IR*_0_ *− IR*_*R*_)*/IR*_0_ to compare the effect of the alternative resections between different patients (figure 4C). At the group level, the SF group presented a larger decrease in seizure propagation (*F* (19) = 25.88, *p <* 10^*−*65^), and a larger effect of increasing the resection size (*F* (19) = 2.90, *p* = 4 *·* 10^*−*5^). There were large differences in *δIR*(*R*) as a function of the resection size between different patients. Whereas in the majority of the cases *δIR*(*R*) increased roughly exponentially with *S*(*R*), for several patients there was an abrupt (discontinuous) jump at a given resection size which may be small (spanning only a few nodes) or large. In order to quantify these differences, we defined the *optimal resection R*_*op*_ as the one leading to a 90% decrease in seizure propagation, *δIR*(*R*_*op*_) = 0.90. The SF group had significantly smaller optimal resections (figure 4D-left, diff=*−*4.34, *p* = 0.03, *rks* = 411.5), and these presented a significantly larger overlap with the actual resection strategy *Ov*(*R*_*op*_, *RA*) (figure 4E-left, diff= 0.20, *p* = 0.03, *rks* = 495.5), that the NSF group. We found good classification results using either of these variables to classify between the SF and NSF groups (*AUC* = 0.71, 0.69 respectively for *S*(*R*_*op*_) and *Ov*(*R*_*op*_, *RA*), figure 4D,E). Both variables led to very similar classification results at the optimal classification point (Youden criterion), correctly identifying 6*/*8 NSF cases. In particular, we found accuracy = 0.74, precision = 0.46, sensitivity = 0.75, *F* 1 = 0.57 for *S*(*R*_*op*_), and accuracy = 0.71, precision = 0.43, sensitivity = 0.75, *F* 1 = 0.55 for *Ov*(*R*_*op*_, *RA*). The classification results for both the validation and modeling cohorts are summarized in table 1.

**Table 1:**
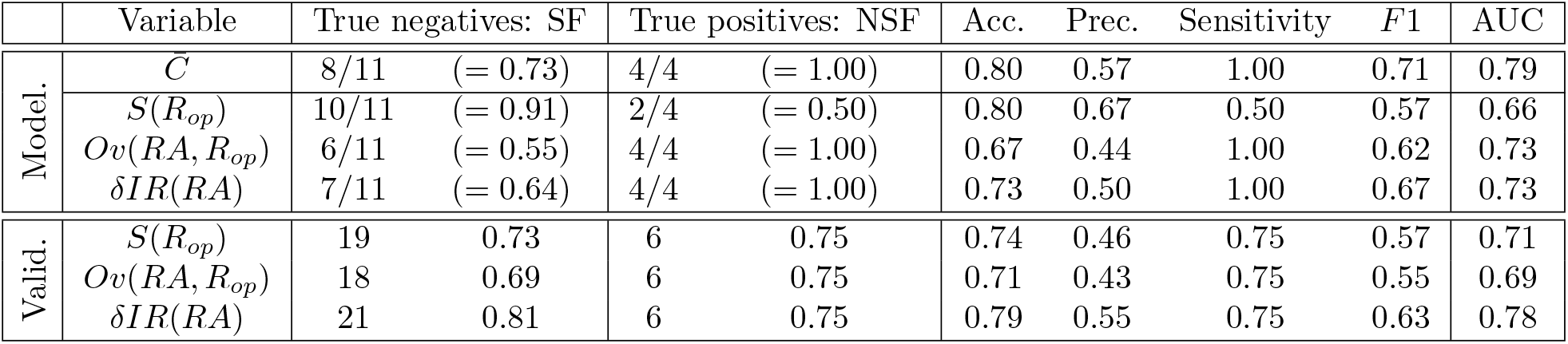
Results of the classification analyses for the modeling (Model.) and validation (Valid.) cohorts. Results correspond to the optimal points of the ROC curves according to the Youden criterion to account for class imbalance. For each group (SF, NSF), we show the number of correctly identified cases by absolute number and relative frequency. The remaining columns correspond respectively to the accuracy (Acc.), precision (Prec.), sensitivity, F1 statistic and area under the curve (AUC).

In summary, these results indicate that the planned resection strategy (accounted for here by the resection area) presented a larger overlap with the optimal resection for patients with good outcome. In particular, there was only a 90.0% of SF and 42.9% of NSF patients were correctly classified by *Ov*(*RA, R*_*op*_). Remarkably, ESSES could also distinguish between SF and NSF patients without taking into account the information of the surgical plan. In fact, up to 90.4% of SF and 46% of NSF patients were correctly identified by *S*(*R*_*op*_) (in relation to only a 76.5% SF-chance and 23.5% NSF-chance according to the group ratios). As this analysis did not depend on the planned resection strategy, a bad prognosis according would be indicative of the need to perform a more exhaustive presurgical evaluation, and potentially imply an unavoidable non-seizure free outcome after any surgery.

Finally, we note that almost equivalent results may be obtained by considering the disconnecting resection, i.e. the smallest resection leading to disconnection of the seed, instead of the optimal resection (see Supp. section 5.4 and Supp. figure 5). This is due to the strong link between network topology and emergent SIR dynamics, a result that can be used to speed up computations considerably, by using a purely network-based analysis of the effect of different resection strategies.

### 2.4 Simulation of the surgical plan

We simulated the effect of the planned surgery in ESSES for each patient by performing virtual resections of the resection area, which was considered as a proxy for the surgical plan here. We report here on the results for the validation cohort (figure 5), results for the modeling cohort can be found in the supplementary information (Supp. section 6, Supp. figure 6). As in the previous section, all modeling details had already been set during the modeling step. The effect of the resection strategy on (modeled) seizure propagation, *δIR*(*RA*), was significantly larger for the SF than the NSF group (figure 5B, *δIR*(*RA, SF*) *− δIR*(*RA, NSF*) = 0.26, *rks* = 513, *p* = 0.02). A ROC classification analysis revealed a good classification between the two groups (*AUC* = 0.78, figure 5C) and at the optimal point (Youden criterion black asterisk in panel C) the majority of patients were correctly identified (accuracy = 0.79, precision = 0.55, sensitivity = 0.75, *F* 1 = 0.63, figure 5D). In particular, there was a 91.3% chance that a patient classified as SF had a good outcome, and a 54.5% chance that a patient classified as NSF had a bad outcome, compared to a 76.5% and 23.5% chance based on the relative group sizes.

**Figure 5:**
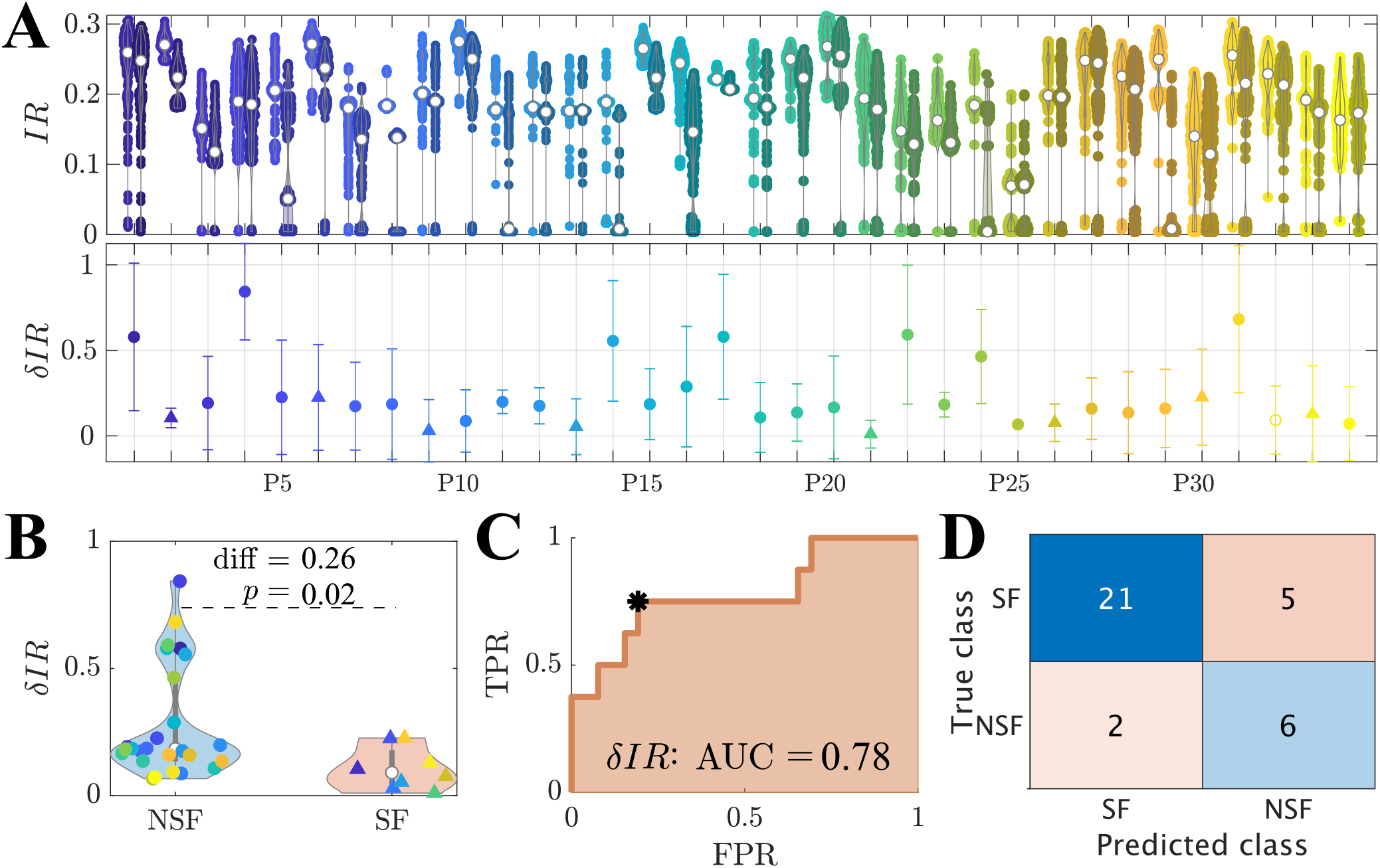
Simulation of the planned resection strategy (validation cohort). **A** The top panel shows seizure propagation *IR* before (left point cloud for each patient) and after (right point clouds) the resection, for 300 iterations of the seed regions, for each patient. The bottom panel shows the average relative decrease in seizure propagation *δIR*(*RA*), with errorbars given by the standard deviation over seed iterations. **B** Comparison of the relative decrease in seizure propagation *δIR*(*RA*) between the SF and NSF groups. Each point corresponds to one patient. **C** ROC curve of the group classification based on *δIR*(*RA*). TPR and FPR indicate respectively the true positive (NSF cases classified as NSF) and false positive (SF cases classified as NSF) rates. **D** Classification results for the optimal point (black asterisk in panel B) of the ROC curve according to the Youden criterion.

### 2.5 Prediction of surgical outcome

The classification analyses in the previous sections were informed by each patient’s surgical outcome. In a prospective setting the outcome for the patient is not yet known, and thus cannot be used to build the classification model. In order to emulate a true prospective setting, we performed a *prediction analysis* based on leave-one-out crossvalidation. That is, in order to predict the outcome of each patient of the validation cohort, a prediction model was built using data from the remaining 33 cases. The results from these analyses were slightly worse than in the previous section, particularly for the NSF class where there was a 12.5% reduction in the group size. In any case, respectively 4, 5 and 5 NSF cases and 19, 18 and 21 SF cases were correctly identified by each ESSES biomarker (figure 6A). Moreover, 75% of NSF cases (6*/*8) and only 19.2% (5*/*26) of SF cases were identified by two or more biomarkers as NSF (figure 7B). For this cohort, if ESSES predicted a good outcome with at least two markers, there was a 80.8% chance of seizure freedom after the surgery (compared to a 76.5% expectancy of surgery success according to the group rates). Conversely, if the model predicted a bad outcome, then there was a 75% chance that the surgery would fail (compared to a 23.5% expectancy of surgery failure according to the group rates). In clinical practice, a good ESSES prediction could then be interpreted as a large (80.8%) chance of seizure freedom after the surgery and thus support the decision to proceed with surgery. On the contrary, a bad ESSES prediction would indicate a 76.5% chance that the surgery would fail. This may be suggestive of the need of more presurgical evaluations or a different resection strategy, and eventually indicate a low probabily of complete seizure freedom after the surgery.

**Figure 6:**
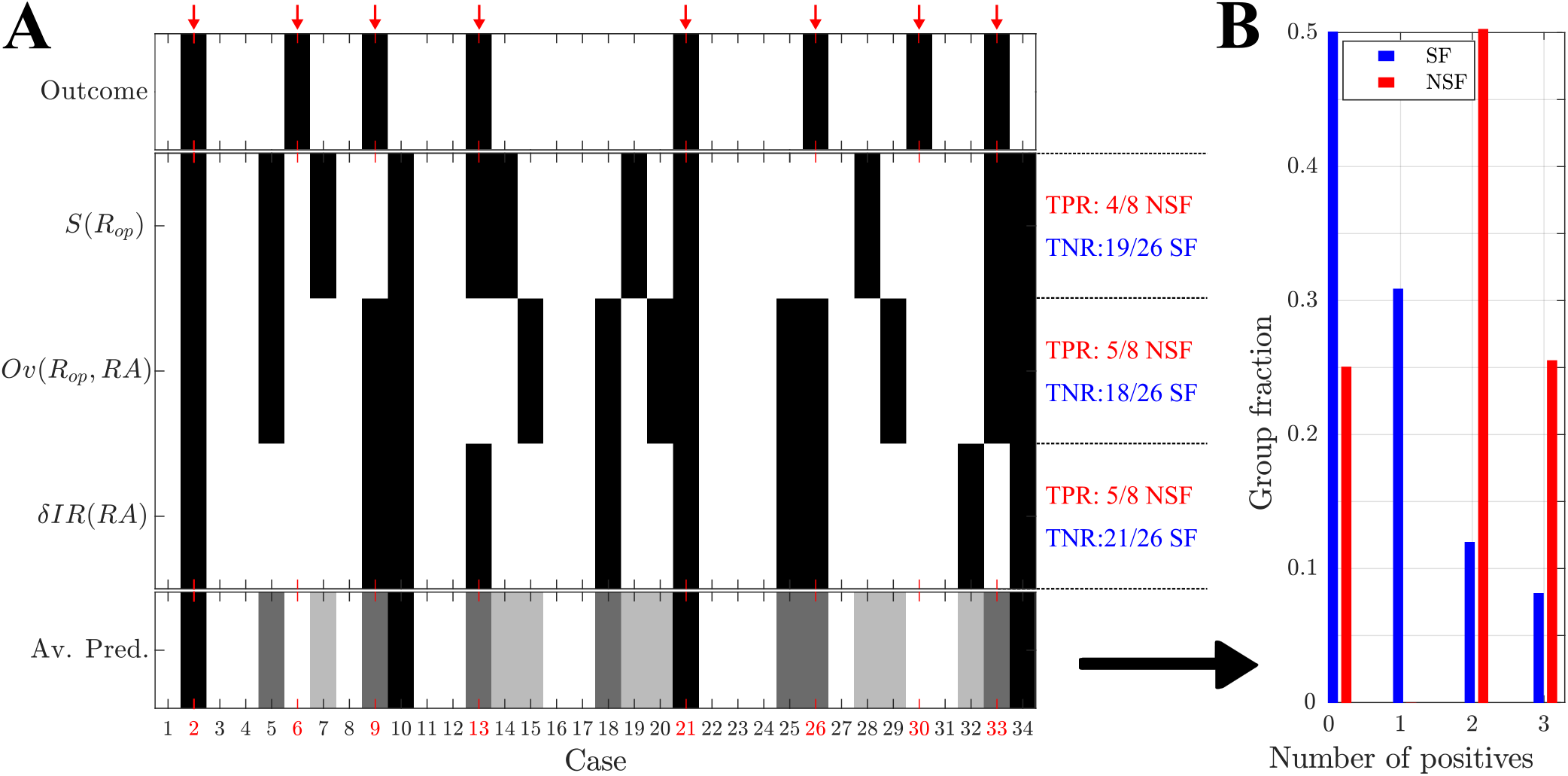
Prediction of surgical outcome: pseudo-prospective study. **A** Prediction results using each of the three model-based biomarkers of surgical outcome: the size of optimal resections *S*(*R*_*op*_), the overlap between optimal resections and the resection area, *Ov*(*R*_*op*_, *RA*), and the decrease in seizure propagation due to simulation of the planned resection strategy, *δIR*(*RA*). NSF (SF) cases are shown by black (white) rectangles. The bottom row shows the fraction of models (0 *−* 3 out of 3) with a positive (i.e. NSF) classification, for each patient. Surgical outcome is shown in the top row. NSF cases are highlighted by a red arrow and by red labels. **B** Relative number of cases identified as NSF by *n* models, *n* = 0, 1, 2, 3, respectively for the SF (blue, left-side bars, *N* = 26) and NSF (red, right-side triangles, *N* = 8) groups.

**Figure 7:**
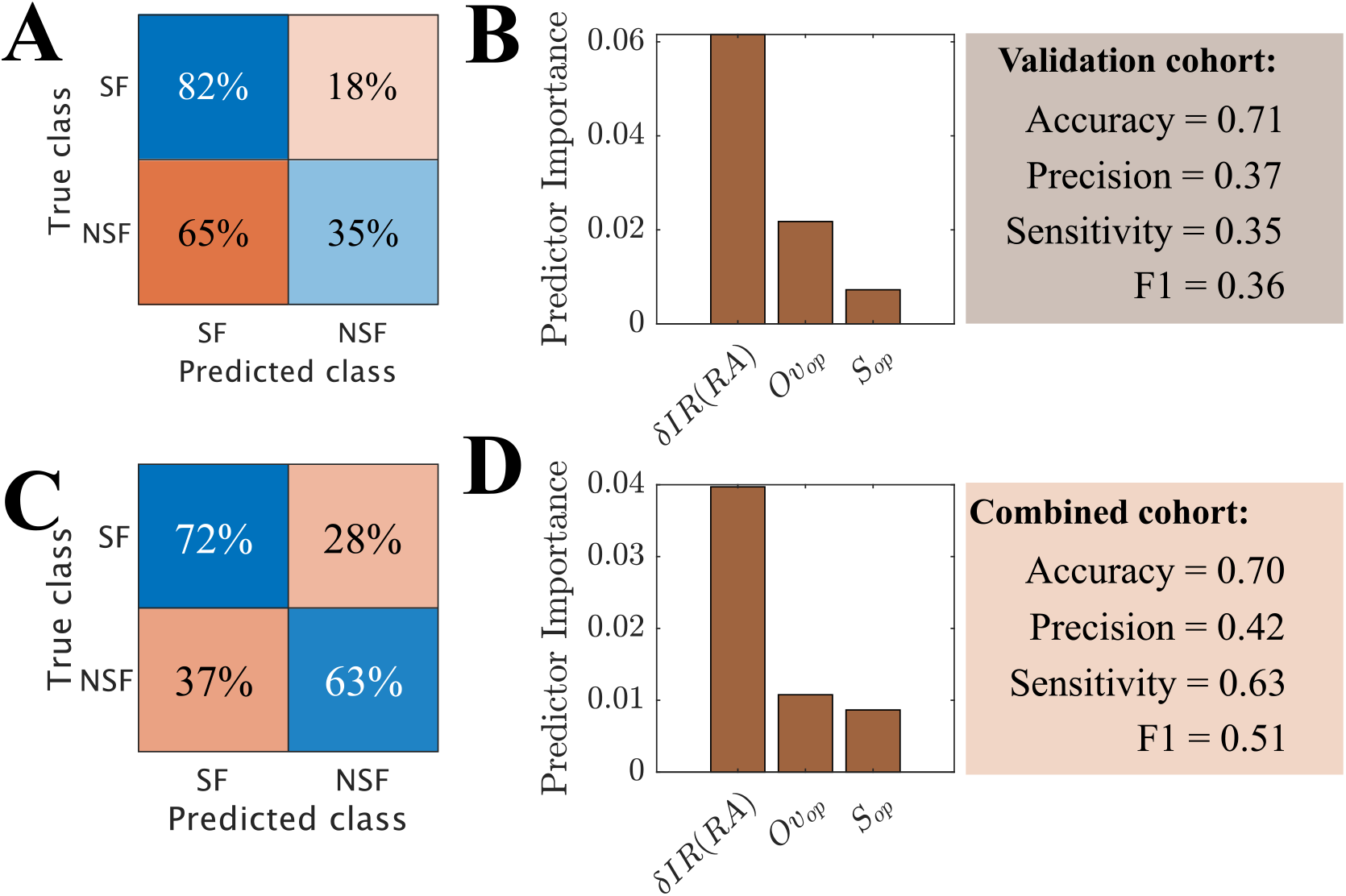
Prediction of surgical outcome using a machine learning algorithm (RUSBoost) with leave-one-out cross validation. As input variables we used the normalized decrease in seizure propagation after virtual resection of the RA, *δIR*(*RA*), the size of optimal resections *S*(*R*_*op*_) and the overlap of optimal and clinical resections *Ov*(*R*_*op*_, *RA*). Panels **A**,**B** show the confusion matrix and predictor importance for the validation cohort (*N* = 34, 8 NSF), and panels **C**,**D** are for the combined cohort (*N* = 49, 12 NSF).

Finally, in order to test whether the information provided by the three biomarkers could be combined to improve the prediction results, we performed a machine learning analysis using an adaptive boosting algorithm with random undersampling and leave-one-out cross-validation (figure 7A,B). The input variables for the classification algorithm where *δIR*(*RA*), *S*(*R*_*op*_) and *Ov*(*RA, R*_*op*_). We found that, even though the accuracy of the model was good (0.71) the machine learning model was biased towards the majority class (SF), with only 35% of NSF cases correctly identified (precision = 0.37, sensitivity = 0.35) and a poor result for *F* 1 = 0.36, even though the considered algorithm (RUSboost) was designed to correct for class imbalance. However, the minority class in our case contained only 8 cases, likely preventing the model from being able to generalize. In order to address this issue, we created a *combined cohort* (*N* = 49) pooling together the patients from the modeling and validation cohorts (figure 7C,D). The combined cohort had 12 NSF cases (50% increase), and the new model was able to identify the majority of NSF cases correctly (72% of SF cases and 63% of NSF cases). Even though the accuracy of the model (0.70) did not improve, the remaining measures, which are less affected by class imbalance, did (precision = 0.42, sensitivity = 0.63, *F* 1 = 0.51). Overall, the machine learning model was not able to improve upon the results found using the individual variables (see table 2). Due to the small sample size, we could not determine whether this was due to intrinsic model limitations, suboptimal hyperparameters, or simply a too small group size (particularly of the minority class).

**Table 2:** Results of the predictive analyses for the validation and combined cohorts. For each analysis, we used a leave-one-out crossvalidation such that a predictive model was build to predict the outcome of each patient using the data from the remaining *N −* 1 patients. For the individual variables, the results correspond to the optimal points of the ROC curves according to the Youden criterion. For the machine learning analyses, they were derived from an adaptive boosting (AdaBoost1, Matlab 2018) algorithm with leave-one-out crossvalidation, combined with random undersampling (RUSboost) to account for class imbalance. Results were averaged over 10 iterations of the AdaBoost1 algorithm. For the combined method, the results from the three individual analyses were combined, and a NSF classification was assigned to patients with at least to positive (NSF) classifications. For each group (SF, NSF), we show the number of correctly identified cases by absolute number and relative frequency. The remaining columns correspond respectively to the accuracy (Acc.), precision (Prec.), sensitivity and F1 statistic. For machine learning analyses only the average fraction of correctly predicted cases is shown in the true negatives and true positives columns, since absolute results can vary per realization of the prediction algorithm.

|  | Variable | True negatives: SF |  | True positives: NSF |  | Acc. | Prec. | Sensitivity | F1 |
| --- | --- | --- | --- | --- | --- | --- | --- | --- | --- |
| Validation | $S(R_{op})$ | 19/26 | (= 0.73) | 4/8 | (= 0.50) | 0.68 | 0.26 | 0.50 | 0.38 |
| | $OV(RA, R_{op})$ | 18/26 | (= 0.69) | 5/8 | (= 0.63) | 0.68 | 0.38 | 0.63 | 0.51 |
| | $\delta IR(RA)$ | 21/26 | (= 0.81) | 5/8 | (= 0.63) | 0.76 | 0.50 | 0.63 | 0.57 |
|  | Combined | 21/26 | (= 0.81) | 6/8 | (= 0.75) | 0.79 | 0.55 | 0.75 | 0.65 |
|  | RUSboost | 0.82 |  | 0.35 |  | 0.71 | 0.37 | 0.35 | 0.36 |
| Combined | RUSboost | 0.72 |  | 0.63 |  | 0.70 | 0.42 | 0.63 | 0.51 |

## 3 Discussion

Personalized models of brain dynamics can aid the treatment of patients with neurological disorders. In this study we presented ESSES (Epidemic Spreading Seizure and Epilepsy Surgery model): a framework to aid epilepsy surgery planning in a patient-by-patient basis. ESSES defines individualized seizure propagation models that integrate multimodal presurgical data, and can propose alternative resection strategies and provide confidence bounds for the probability of success of a given strategy. The implementation of ESSES in clinical practice may thus eventually improve the chances of achieving a good postsurgical outcome.

In this study we proposed a combined setting such that ESSES’ parameters were fitted on a retrospective study (*N* = 15) using iEEG data of ictal activity, in analogy with previous studies [19, 21, 27, 38, 40, 41]. We validated that ESSES captured the main aspects of seizure propagation and was able to reproduce the iEEG-recorded seizures, in agreement with our previous studies [30, 32]. Remarkably, the goodness-of-fit of the ESSES-modeled seizures to the iEEG data could identify patients with a bad outcome with *AUC* = 0.79, 100% sensitivity and 57% precision. Such information may be integrated in the presurgical evaluation of the patients for whom iEEG data is available: different resection strategies may be tested as the origin of the ESSES-modeled seizures [32], with a low goodness-of-fit being indicative of a low chance of seizure freedom. In particular, a bad prediction by the model would indicate (in this cohort) a 57% chance of a bad outcome (to be compared with only a 26.7% NSF rate in this cohort). Conversely, all patients identified as SF by the model could proceed to surgery with high expectations (100% in this group) of good outcome.

The novel aspect of this study consisted of a subsequent *pseudo-prospective study* with an *independent cohort* and in a *blind setting*. Importantly, we did not require the presence of iEEG data on the pseudo-prospective study, and instead the multimodal presurgical information available for each patient was integrated into seed-probability maps. In this manner ESSES can be adapted to the information available for each patient, in a quantitative and systematic manner. IEEG data is highly invasive and burdensome for the patient, and thus not always part of the presurgical evaluation. For instance, only 19 of the 34 patients of the validation cohort had undergone it. Thus, by not requiring iEEG data the ESSES framework can be applied to a much larger patient population than traditional approaches [19, 21, 40], with the expected wider impact.

ESSES may be applied prospectively as follows. First of all, ESSES may suggest optimal resection strategies, in analogy with previous studies [23, 24, 30, 42], with the advantage that all multi-modal presurgical information available for each patient is integrated into ESSES, instead of considering only one source used for network reconstruction. The *optimal resection strategy*, defined here as the smallest resection leading to a 90% decrease in (modeled) seizure propagation, can be used as a first indicator of the chances of seizure-freedom after *any* surgery. In our pseudo-prospective predictive framework (emulating the presurgical conditions) the size of this resection could predict 50% of patients with bad outcome (table 2), whereas the relative NSF rate in this group was 23.5%. This result is independent of the resection strategy and it is completely characterized by the presurgical information available for each patient. Thus, a bad prognosis could indicate that either the presurgical information available is not of sufficient quality, or that the patient is unlikely to be seizure-free with any resection strategy.

ESSES can also provide information about the prognosis after a particular resection by i) comparing it to the optimal ESSES resection strategy and ii) quantifying its effect on seizure propagation in the patient-specific ESSES model. Here we found that resections with a larger overlap with the optimal resection were more likely to lead to seizure freedom, in agreement with previous studies [19, 27, 40]. Similarly, resections leading to a larger decrease in seizure propagation in ESSES were associated with a larger probability of seizure-freedom after the resection, in agreement with other modeling [19, 40] and network-based [7, 12, 20] studies. Here we considered only the planned resection strategy, which was approximated here by the resection area, since this information could be derived in a systematic manner, and this set-up allowed us to validate ESSES’ findings. In a presurgical setting, different strategies could be tested to measure the probability of seizure freedom after each one. In particular, we found that, when combining the information from the three model-based biomarkers (namely the size of the optimal resection, its overlap with the planned resections, and the effect of the planned resection on modeled seizure propagation) could predict pseudo-prospectively 81% and 75% of SF and NSF cases (see table 2), whereas the relative group ratios were 76.5% and 23.5%, respectively. Clinically, this implies that if a good prognosis is found by at least two biomarkers, then there is a 91.3% (true negative rate, 21 cases were SF of the 23 predicted by the model) chance that the patient will be seizure-free, and the patient can proceed with the surgery with the knowledge that they will likely have a good outcome. Conversely, a bad prognosis by at least two biomarkers indicates a 55% chance of bad outcome, and may be interpreted as an ESSES suggestion to perform more presurgical testing or consider alternative resection strategies. Importantly, epilepsy surgery may still improve the quality of life of the patient even in chases for which complete seizure freedom cannot be achieved. Thus, moderate *a priori* chance of a bad outcome is not necessarily a contraindication for surgery but it is important in the presurgical counseling of the patients.

Our findings here did not depend on the presence of iEEG data, and even iEEG data were available we only included a low-resolution description of them. IEEG data does provide the most detailed information of epileptogenic activity, and is it often the most valuable tool to identify the epileptogenic zone or predict surginal outcome for patients with complicated ethiology [22, 25, 27, 33–36]. In fact, for the modeling cohort we found the best classification results using the goodness-of-fit of the ESSES-predicted seizure propagation patterns to the iEEG seizures, in agreement with previous studies [27]. IEEG imaging however is burdensome to the patient, has risk of complications, and has limited spatial coverage. A first prediction of surgical outcome could thus be performed with the ESSES framework when the results of non-invasive testing have been obtained, and an iEEG study might be avoided if the model already predicts a good outcome with the existing data.

In summary, we showed here that ESSES could identify patients with good outcome presurgically based on i) the smaller size of the optimal ESSES resection strategies, ii) a larger overlap of the planned resection strategy with the optimal ESSES resection, and iii) a larger effect of the planned resection strategy on decreasing (modeled) seizure propagation. Our findings here indicated that ESSES could be generalized to other patient populations (as we did with the validation cohort), with the only requirement of a patient-specific brain network, and can incorporate multimodal information from the existing presurgical evaluation, in particular without requiring the presence of iEEG data. The ESSES-based biomarkers identified here could be taken into account during presurgical planning to evaluate the need for more testing, or may lead to the decision to forgo the surgery, if a bad outcome is predicted. This extra information may be particularly valuable for patients with complicated ethiology (e.g. discordant information from different modalities, variable seizure propagation patterns, multiple seizure onset zones), for whom the discussion of whether or not to perform the surgery is challenging.

### 3.1 ESSES modeling framework

ESSES consists of different interconnected elements, namely i) the underlying network structure; ii) the seizure propagation model (and parameter fitting); iii) the seizure onset zone model; iv) the virtual resection model; and v) the virtual resection optimization model. Each of these different elements was designed to model a particular aspect of epilepsy surgery in a synergistic manner. For instance, the emergent properties of the seizure propagation model (the SIR model) led the design of the virtual resection optimization algorithm. At the same time, the modular organization of the framework allows for the independent improvement or modification of each of the modules. In fact, different modules were developed and analyzed in detail in our previous studies. For instance, the virtual resection algorithm model was initially designed in Ref. [24] and improved in Ref. [30], whereas the seizure propagation and parameter fitting model as used here was mainly defined in Ref. [32]. Below we discuss the main modeling considerations and results for each ESSES module.

As the underlying network structure we considered MEG-derived whole-brain networks as a proxy for structural connectivity, following our previous works [30, 32], and in contrast with other works [21, 24, 29, 42]. MEG provides highly temporally resolved information with good spatial resolution and uniform coverage. Our previous studies showed that MEG networks based on the amplitude envelope correlation (AEC) can integrate information from both short-range structural connections (by not correcting for volume conduction) and long-range functional coupling. Thus, AEC-MEG networks can be used as a cost-effective proxy for structural connectivity [30] with much lower computational cost than DTI networks, whilst also being more sensitive to long range connections, in particular inter-hemispheric ones, that may often be missed by DTI [43].

ESSES was based on a simple epidemic spreading model, the SIR model. Epidemic spreading models, such as the SIR or SIS (Susceptible-Infected-Susceptible) models, describe the basic aspects of spreading phenomena on networked systems [31], and have been used to describe other neuro-physiological processes before, such as the spreading of pathological proteins on brain networks [44, 45] or the relation between brain structure and function [46]. Epidemic spreading models have been extensively studied on different network substrates [31] and are supported by a well-grounded mathematical and computational framework that we can use to our advantage in the context of epilepsy surgery. For instance, from an epidemic spreading perspective, it is to be expected that hub removal plays a major role in the decrease of seizure propagation, as found experimentally [12, 20], with the spreading threshold heavily influenced by the existence of hubs [31]. This theoretical background guided the design of an efficient virtual resection optimization algorithm, such that the decrease in seizure propagation after a virtual resection could be approximated by the decrease of centrality of the seed regions.

As we showed here and in previous works, epidemic spreading models can also reproduce the fundamental aspects of seizure propagation at the whole-brain level in epilepsy patients [30, 32]. As the working point of the ESSES model we chose here the values of the global parameters that led to the maximum average goodness-of-fit of the modeling cohort (figure 1). Importantly, ESSES was still individualized for each patient by means of the patient-specific brain connectivity, setting the local spreading probabilities, and the patient-specific seed regions (based on the seed-probability maps built with multi-modal presurgical information). As we showed in our previous study [32] and in the supplementary information here (Supp. section 5.2), by not individualizing the global model parameters (namely *ρ* and *γ*) for each patient we were able to reduce noise effects by integrating together ictal data from different patients. Moreover, this formulation allowed us to generalize ESSES to patients for whom iEEG seizure-propagation patterns were not available.

Our findings in this study indicated that the iEEG seizure propagation patterns were significantly better explained by ESSES for SF patients, and in fact all NSF cases could by identified by a bad ESSES fit, and 73% of the SF cases by a good fit. There are several possible explanations for these findings. Given that the epidemic seed was based on the resection area for each patient in this part of the analyses, a simple explanation is that the resection strategy might have been better for SF patients given the existing information. However, the difference could also arise from the iEEG data: the sampling may have been inadequate for NSF patients [29], or these may have presented seizure *dynamotypes* [47] that were not well-explained by the considered epidemic spreading model (SIR model). The fact that the optimization of virtual resections analysis –which did not depend on the clinical resection area– also found differences between the SF and NSF groups points towards and intrinsic difference between the presurgical data of the two groups, and not only to a sub-optimal surgical strategy for the NSF group. The next ingredient of ESSES was the definition of the seizure onset zone in the model, that is, the set of brain regions from which seizures originate. In this study we presented a method to combine the multimodal presurgical information available for each patient into *seed-probability*. This set-up thus emulated the clinical situation prior to the surgery, where a surgical strategy has been devised based on the available information from the presurgical evaluation. It would also allow for flexibility in the clinical application of ESSES: if more evaluations become available these could be readily integrated into the seed-probability map to update ESSES’s results.

The final key ingredients of ESSES were the simulation and optimization of resection strategies. Here we considered a node-based resection such that the resected nodes were disconnected from the network. This approach however does not take into account possible widespread effects or plasticity mechanisms, which could also be included into the model [48]. The virtual resection optimization algorithm was originally validated in our previous studies [24, 30]. Given that optimizing virtual resections is highly computationally demanding, the algorithm took advantage of the mathematical link between network structure and SIR dynamics to reduce the dynamics-based optimization problem (i.e. finding the resection leading to a minimum seizure propagation) into a network optimization problem (i.e. finding the resection leading to a minimum seed efficiency). This was also motivated by our previous finding that the effect of a resection on the model depended strongly on the centrality of the seed regions after the resection [24, 30]. In particular, Nissen et al. [24] found that removing connections to the network hubs was the most efficient way to decrease seizure propagation, whereas Millán et al. [30] verified a strong correlation between a decrease in closeness centrality of the seed and a decrease in seizure propagation following a virtual resection. The effect of a resection on seizure propagation is also influenced by other network and model properties, and as a consequence the optimal network-based and SIR-based resections may differ slightly [30]. However, the intrinsic noise in the seed definition, in the seed-probability maps, and in the actual origin and propagation patterns of iEEG-recorded seizure created variability in the clinical data that absorbed the differences between the network-based and SIR-based optimal resections (which we previously found to be small anyway [30]). The virtual resection optimization algorithm considered here imposed no conditions on the location of the resected regions, nor did it force that the resection strategy was made up of only one set of adjacent regions. Conditions on the resection strategies could be imposed, such as preserving eloquent cortex or forbidding bi-hemispheric resections [23, 42]. This would limit the dimensionality of the space of possible resection strategies and simplify the computations. However, by not imposing any conditions here we derived an *optimal* ESSES resection against which other, perhaps clinically more realistic, strategies could be tested (by e.g. measuring their overlap as we did here).

### 3.2 Modeling considerations and limitations

There are inherent limitations in the modeling of virtual resections, as the findings cannot be directly tested and we often rely on retrospective data. Here we have attempted to simulate how an epilepsy surgery model could be used in the clinic, i.e. prospectively, by considering only the presurgical information that is typically available to the clinical team. However, the optimal resections suggested by ESSES can still not be tested in practice. Only long-term testing of the framework in the clinic can truly validate the use of computational models in epilepsy surgery.

ESSES is an abstraction of seizure dynamics that does not aim to reproduce the detailed bio-physiological processes involved in seizure generation and propagation, but aims to focus only on the most relevant features of seizure propagation [24, 29, 30, 32]. In order to validate ESSES as a framework to simulate seizures, we compared the modeled seizures with those recorded via iEEG. This required however of a simplified representation of the iEEG data. In particular, as there was no intrinsic time-scale in the SIR model, and to avoid introducing an arbitrary one, we reduced the iEEG data to a pattern that describes the activation order of the sampled ROIs. Furthermore, even if ESSES provides a good representation of the iEEG seizures, extrapolating these results to the simulation of the effect of a resection is not trivial. Moreover, our virtual resection technique assumed that the effect of a surgery could be approximated simply by removing or disconnecting the resected regions, whereas in practice widespread effects and compensation mechanisms are expected [48]. Here we validated ESSES’ results against postsurgical outcome, but seizure freedom is not a perfect gold standard either. For instance, in cases with a good outcome a smaller resection could potentially also have led to seizure freedom [24, 30].

All modeling frameworks are affected by the need to (sometimes arbitrarily) choose modeling parameters, which go from the data reduction process to the choices of thresholds and metrics for the final analyses. Here we considered well-established data preprocessing techniques [49]. ESSES was validated in previous studies [24, 30, 32], and importantly we found that the results held for an independent cohort, and that modeling details (such as the simulation algorithm for the SIR model) did not affect the main results [32]. A simple model to simulate seizure propagation (the SIR model), also reduced the number of modeling parameters so that the findings could be more easily generalized. Some arbitrary choices were still needed, such as the definition of the 90% threshold to select the optimal resection strategy. However we validated that similar results were obtained when another resection (the disconnecting resection) was considered.

The seed-probability maps were based on an existing low-resolution database [39]. Seed regions were consequently widespread over the network. This also led to large variability in the analyses for each patient (see for instance figures 4A-C and 5A), as the results of each simulation depended strongly on the seed realization. In order to improve the resolution of the model and minimize noise, the data from each modality could be integrated directly into the model, skipping the database 34-region description.

Finally, it is a limitation of this study the small size of the non-seizure-free group, with only 4 cases in the modeling cohort and 8 in the validation cohort. This small size limited the classification and prediction analyses, and prevented us from building a more sophisticated machine learning model based on our analysis. Future studies involving more that one center have the potential to at least diminish this limitation.

## 4 Conclusion and outlook

Individualized computational models of seizure propagation and epilepsy surgery based on patient-specific brain connectivity can reproduce individual iEEG seizure propagation patterns and aid epilepsy surgery planning by proposing alternative resection strategies and providing estimates on the likelihood of seizure freedom after the surgery. Here we presented the ESSES framework for seizure propagation and epilepsy surgery. ESSES combines SIR epidemic spreading dynamics over patient-specific MEG brain connectivity with a virtual resection framework. We defined a method to derive patient-specific regional epileptogenicity maps from the presurgical evaluations of the patients in a systematic and quantitative manner, and integrated them into ESSES. We performed a pseudo-prospective study emulating the use of ESSES in clinical practice, prior to surgery. In the pseudo-prospective analyses we did not require the presence of iEEG data, demonstrating that the model could be applied to larger patient populations. We found that the goodness-of-fit of ESSES to the iEEG seizures (in a retrospective study), the effect of the planned resection strategy, as well as the size of ESSES optimal resections and their overlap with the planned resection predicted surgical outcome with 0.68 *−* 0.76 AUC and 0.50 *−* 0.63 sensitivity to identify non-seizure-free patients. Our results thus prescribe the use of ESSES during the presurgical evaluation to evaluate the need for further presurgical testing on a case-by-case basis or, conversely, support the decision to proceed with surgery in the case of a good-outcome prediction. For cases where a bad outcome is predicted, the surgical plan may be altered to include the ESSES information.

## 5 Methods

The general design of the study is detailed in figure 2. Namely, we first set the hyperparameters of ESSES using a *modeling cohort* (*N* = 15) for which seizure propagation patterns derived from iEEG recordings were available. Then, ESSES was fitted with multimodal patient-specific data (in the form of seed-probability maps), and it was used to a) identify optimal resection strategies for each patient and b) predict the chance of a good outcome after a given resection. Then, ESSES was applied to a *validation cohort* (*N* = 34) in a pseudo-prospective analysis with a blind setting to emulate the presurgical conditions. That is, during the application of ESSES to determine optimal resection strategies, the researchers were blind to the actual clinical resection and surgery outcome of each patient. This data was subsequently de-blinded in two stages. First, the resection areas were obtained to be used as a proxy for the surgical plan of each patient to a) compare them with the ESSES optimal resection strategy, and b) simulate the effect of the surgical plan in the ESSES framework. Finally, we de-blinded the one-year surgical outcome to enable a statistical validation of the results.

### 5.1 Patient groups

We included two patient groups in this study, the *modeling cohort* for the model definition (retrospective study) and the *validation cohort* for the pseudo-prospective validation. All patients had undergone resective surgery for epilepsy at the Amsterdam University Medical Center, location VUmc, between 2013 and 2019. All patients had received an MEG recording, and underwent pre-and post-surgical magnetic resonance imaging (MRI). All patients gave written informed consent and the study was performed in accordance with the Declaration of Helsinki and approved by the VUmc Medical Ethics Committee. The excluding criterion was the existence of a prior brain surgery.

Both patient groups were heterogeneous with temporal and extratemporal resection locations and different etiology (see Supp. tables 1 and 2 for details). Surgical outcome was classified according to the Engel classification at least one year after the surgery [50]. Patients with Engel class 1A were labelled as seizure-free (SF), and patients with any other class were labelled as non-seizure-free (NSF). The modeling cohort consisted of 15 patients (4 NSF, 11 females) who had also undergone an iEEG (invasive electroencephalography) study, including post-implantation CT-scans. This same cohort was already included in Ref. [32], and partially in [30]. The validation cohort consisted of 34 patients (8 NSF, 13 females). No extra requirements (other than the presence of an MEG recording of sufficient quality) were placed. In order to maintain the pseudo-prospective setting, the research team was blind to the resection area and outcome of the validation cohort patients. In order to perform the final analyses, for which this information was needed, the data was coded to avoid identification. For two cases of the validation cohort (cases 2 and 9) the data of surgical outcome was de-blinded together with the data of the resection area as the research team became aware of a subsequent resective surgery (indicative of a bad outcome of the first surgery).

### 5.2 Individualized Brain Networks

Seizure propagation was modeled on the patient-specific brain networks, as derived from MEG data, for both cohorts (see figure 2). For each patient, a 10 to 15 minutes eyes-closed resting-state (supine position) MEG recording was used to derive broadband (0.5 -48.0 Hz) MEG functional connectivity. All instrumental and methodological details were equal to our previous studies [30, 32] and are detailed in the supplementary information (Supp. section 2). Functional networks were generated considering each of the 246 ROIs of the Brainnetome (BNA) atlas [51] as nodes. The elements *w*_*ij*_ of the connectivity matrix, indicating the strength of the connection between ROIs *i* and *j*, were estimated by the AEC (Amplitude Envelop Correlation) [52–55], without including a correction for volume conduction. The uncorrected AEC maintains information about the structural connections, which are mainly determined by the distance between each ROI pair, by not correcting for volume conduction, it. We validated the relationship between AEC-MEG and structural networks in a previous study [30] by comparing them with a well-validated model for structural connectivity: the exponential distance rule (EDR) network. Based on animal studies, the EDR specifies that the weights of structural connections in the brain, *w*_*ij*_, decay exponentially with the distance between the ROIs *d*_*ij*_ [56–58], i.e. *w*_*ij*_ *∝ exp*(*−αd*_*ij*_). Recent studies have corroborated that the EDR reproduces human DTI data well [59–61], and in Ref. [30] we validated that AEC-MEG networks were strongly correlated (*R*^2^ = 0.50) with the corresponding EDR networks, therefore showing that AEC-MEG encodes structural information. Moreover, AEC-MEG networks also include long-range connections that may promote seizure propagation, but that may be missing from structural (i.e. DTI) networks [62, 63]. Thus, uncorrected AEC-MEG networks are a convenient way to construct a network that resembles a structural network and includes long-range connections.

AEC values were re-scaled between 0 (perfect anti-correlation) and 1 (perfect correlation), with 0.5 indicating no coupling [64]. Functional networks were thresholded at different network densities *ρ* indicating the fraction of links remaining in the network. We note that the networks were thresholded but not binearized, so that *w*_*ij*_ could take values between 0 and 1.

### 5.3 Resection Area

The resection area (RA) was determined from the three-month post-operative MRI. For the modeling cohort the resection areas were obtained as part of two previous studies [30, 32]. For the validation cohort, to maintain a completely blind setting for the first analysis (*Optimization of alternative resections*), the resection areas were obtained during a second pre-processing step, as described in figure 2. Cases 9 and 20 of the validation cohort underwent the post-operative MRI on a different MRI scanner at their resection center, respectively one day and three weeks after the surgery. Case 9 also lacked a 3-month postoperative MRI, an MRI from 2 years after the surgery was used instead.

The post-resection MRIs were co-registered to the pre-operative MRI using FSP FLIRT (version 4.1.6) 12 parameter affine transformation. The resection area was then visually identified and assigned to the corresponding BNA ROIs, namely those for which the centroid had been removed during surgery.

### 5.4 iEEG Seizure Propagation Pattern

Patients in the modeling cohort underwent invasive EEG recordings using stereo-tactic electrode implantation as described in [32]. One characteristic iEEG-recorded seizure from each patient was used to derive a seizure propagation pattern in terms on the BNA ROIs, the *iEEG seizure pattern*, as described in [32] and in the Supp. section 3.

### 5.5 Seizure Propagation Model

ESSES was based on our previous studies [24, 30, 32] where we showed that simple epidemic spreading models could reproduce the spatio-temporal seizure-propagation patterns derived from invasive EEG recordings, and that they could be used to simulate the effect of different resection strategies *in silico*. ESSES was based on a well-known epidemic spreading model: the Susceptible-Infected-Recovered (SIR) model [31], which was simulated on the patient-specific MEG brain network. The SIR model simulated the propagation of ictal activity from a set of *seed* regions that were set to be infected at the beginning of the simulation to the remaining nodes in the network, and the subsequent recovery of infected nodes. The SIR dynamics were defined by two parameters: the probability *β*_*ij*_ that each infected node *i* propagates the infection to a neighbour *j* (*S → I*), and the probability *γ*_*i*_ that each infected node *i* recovers (*I → R*). For simplicity, we considered here a global recovery probability *γ*_*i*_ = *γ*, and spreading probabilities given by the MEG network connectivity: *β*_*ij*_ = *w*_*ij*_. Thus, the spreading rate was determined by the density of connections in network *ρ*. The two control parameters of the ESSES model are thus the network density *ρ*, and the recovery probability *γ*. Depending on the network structure, the epidemics can show different spatio-temporal spreading profiles described by the probability *p*_*i*_(*t*) that each ROI *i* becomes infected at step *t*.

*ρ* and *γ* were fitted to the iEEG seizure-propagation patterns at the group level. The resection area was set as the seed of epidemic spreading, and an *ESSES seizure propagation pattern* was built that described the set of infected and non-infected ROIs during the SIR-simulated seizures, as well as the order in which infected ROIs became infected. In order to take into account the stochastic nature of the SIR dynamics, the participation of each ROI was weighted by the fraction of realizations in which it was involved in the simulated seizure (since different ROIs became infected in different realizations). The *goodness-of-fit* of the model, *C*(*ρ, γ*) [32], quantified how similar the ESSES and iEEG patterns were. It took into account two factors: the weighted correlation between activation orders of ROIs that were active in both patterns, *C*_*w*_, and the overlap between the active and inactive ROI sets of both patterns, *P*_overlap_, i.e.

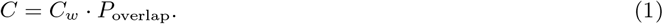

The details of this definition can be found in Supp. section 5.2.

We estimated *C* for a range of values *ρ ∈* {0.025, 0.05, 0.10, 0.20, 0.30} and *γ ∈* 10^*−*4^, 10^*−*3^, 10^*−*2^, 10^*−*1^, considering *N*_*R*_ = 10^4^ iterations of the SIR dynamics 10 times in order to determine average *C* values and their fluctuation for each patient. We then found the parameter set that maximized *C* for each patient (see Supp. section 5.2 and Supp. figure 2) and at the group level (figure 1A). The model parameters that lead to the best fit at the population level defined the ESSES model and were carried over to the pseudo-prospective analyses. Importantly, even though the SIR global parameters were set equal for all patients, ESSES was individualized for each patient by means of their patient-specific MEG brain connectivity, which defined the spreading probabilities, and their patient-specific seed-probability map, which defined the seed regions.

The SIR dynamics was simulated by an adaptive Monte Carlo method (the BKL algoritihm) in Matlab in discrete time, such that at each time step one new node became infected. *N*_*R*_ = 10^4^ iterations of the dynamics were run for each model configuration in all analyses.

### 5.6 Presurgical hypothesis of the seed regions

We built seed-probability maps indicating the probability that each ROI started a seizure, for each patient of both cohorts. This is a key difference with our previous studies, where the seed regions were either derived from the resection area [24, 30, 32], which can only be known after the surgery, or from the iEEG data [30, 32]. Here we defined a framework to integrate data from the different presurgical evaluations that were available for each patient, which was encoded in an existing database (Castor EDC, Ciwit B.V., Amsterdam [39]).

To compute the seed-probability maps, we considered the information available from 6 presurgical modalities: i) presence of ictal activity in EEG, ii) MRI lesions, iii) MEG abnormalities, iv) PET lesions, v) SPECT abnormalities and vi) iEEG recordings of ictal activity. All patients had undergone an EEG, MRI and MEG study, but not all of them presented PET, SPECT or iEEG data. The presence (1) or absence (0) of data of each modality was encoded in a variable *D*_*m*_ = 0, 1, *m* = 1, 2, …, 6, for each patient.

The database included information at the level of 34 regions, consisting of 6 frontal regions (fronto-orbital, frontal-basal, frontal-parasagitaal, frontal-periventricular, frontal-lateral, frontal-operculum), 6 temporal regions (hippocampus, amygdala, uncus, anterior-neocortical, posterior-neocortical, gyrus-parahippocampalis), 2 insular regions (anterior and posterior insula), 1 central, 1 parietal and 1 occipital region, for each hemisphere. The temporal and frontal lobes are the most often involved in EZ and resection strategies, and thus are described in more detail in the database.

For each region *i* and modality *m*, the database indicates the presence (1) or absence (0) of abnormalities, from which we derived binary abnormality maps *a*_*i,m*_ = 0, 1. The overall abnormality map *A*_*i*_ was obtained by aggregating over all modalities available for each patient. Not all modalities are equally relevant to establish the probability that a region is involved in epileptogenic activity: EEG is the least focal, whereas iEEG provides the most localized information, and its results also integrate information from the other modalities (as these affect where the iEEG electrodes are placed). In order to gauge these differences, we weighted each modality *m* by a relevance factor *ω*_*m*_, with *ω* = 1 for EEG, 2 for MRI, MEG, PET and ISPECT, and 4 for iEEG. Thus, the overall abnormality map was defined as

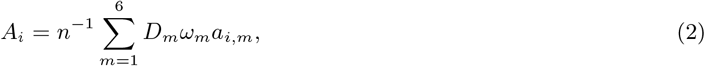

where the normalization factor *n* is defined as 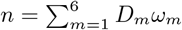

A clinician (ECWvS) defined a unique projection of the regions in the database unto the BNA ROIs. In most cases the database regions corresponded to well-defined gyri that are also well-described in the BNA documentation. A table describing the projection is included as supplementary material. We projected the abnormality map *A*_*i*_ from the low-resolution description into the BNA atlas to obtain the seed-probability maps *SP*_*i*_, with *i* = 1, 2, …, 246. Given that the description provided by the database was broad and homogeneous (i.e. the considered ROIs are much larger than the BNA ROIs), and that co-occurrence of abnormalities in different modalities is a strong indicator of the epileptogenic zone, we included a re-scaling factor *R* to produce more focal seed-probability maps: *SP*_*i*_ = (*A*_*j*_)^*R*^, where *j* is the region in the database corresponding to the BNA ROI *i*. We found that for *R >* 2 the results did not depend strongly on *R*, and report here for *R* = 3.

### 5.7 Virtual Resections

We conducted virtual resections of sets of nodes *R* by disconnecting them from the network, by setting to 0 all their connections. The effect of each resection was characterized by the normalized decrease in seizure propagation *δIR*(*R*) in the resected network (*R*) with respect to the original (0) one:

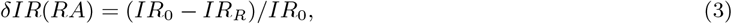

where *IR* is the fraction of nodes that became infected at any point during the modeled seizure, namely,

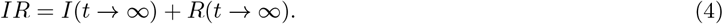

That is, *IR* takes into account all nodes that became infected during the simulated seizure, regardless of whether they eventually recovered or not. This characterizes the total extent of the simulated seizure.

We performed two virtual resection studies, as detailed in figure 2. Firstly, we performed an *Optimization of alternative resections* analysis. We derived optimal virtual resections *R* of increasing sizes *S*(*R*) (defined as the number of resected nodes) with an optimization algorithm based on simulated annealing [65] and derived in our previous studies [24, 30]. The optimization method took advantage of the relationship between SIR spreading and network structure to use a structural metric –the seed efficiency– as a proxy for the actual effect of the resection on seizure propagation *δIR*(*R*). Thus, for each resection size *S*(*R*), the simulated annealing algorithm searched for the resection *R* that minimized the seed efficiency *E*_*R*_(seed) [37, 66, 67]. All nodes were considered as possible targets of the resection. To compare between different patients we defined the normalized seed efficiency

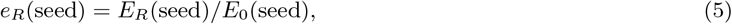

where *E*_0_(seed) is the seed efficiency in the original (un-resected) network. The actual effect of each resection was quantified by the seizure propagation level after the resection, *IR*(*R*), and the normalized decrease in seizure propagation *δIR*(*R*). We defined the *optimal ESSES resection R*_*op*_, as the smallest resection leading to (at least) a 90% decrease in (modeled) seizure propagation. This resection was characterized by its size *S*(*R*_*op*_) and overlap with the resection area *Ov*(*RA, R*_*op*_). We also defined the *disconnecting resection R*_*D*_ as the smallest resection that lead to seed disconnection (see Supp. section 5.4 and Supp. figure 5).

In the second virtual resection study, we simulated the effect of the planned resection for each patient, to measure its effectiveness in reducing seizure propagation. The resection area was used as a proxy for the resection strategy (figure 2: Simulation of the resection plan), since it could be derived in a systematic manner from the data.

For all virtual resection analyses the seed regions were derived from the patient-specific seed-probability maps, and the underlying network was given from the patient-specific MEG network as before. In order to obtain precise results, the effect of each resection was averaged over 300 independent realizations of the seed regions from the seed-probability maps. As described in the right side of figure 2, for the validation cohort we first performed the *Optimization of alternative resections* (figure 2) in a blind setting. Then the resection areas were de-blinded and used as a proxy of the planed resection strategy to i) quantify the overlap of the ESSES optimal resections with the resection strategy and ii) measure the effect of the planed resection in decreasing (modeled) seizure propagation. Finally the one-year postoperative outcome was also de-blinded and used for the statistical analyses.

### 5.8 Statistics

The weighted correlation coefficient was used to determine the correlation between the iEEG and ESSES seizure propagation patterns for the modeling cohort. In all analyses, for comparisons between SF and NSF patients, we used a two-sided Wilcoxon ranksum test. Significance thresholds for statistical comparisons were set at *p <* 0.05.

We performed receiver-operating characteristic (ROC) curve analyses to study the patient classification based on i) the goodness-of-fit of the model (modeling cohort), ii) the size of optimal and disconnecting resections (modeling and validation cohorts), iii) the overlap between optimal resections and the planed resection (modeling and validation cohorts), and iv) the effect of the planed resection on modeled seizure propagation (modeling and validation cohorts). A positive result was defined as bad outcome (non-seizure-free, NSF) classification.

In order to account for the noise in the SIR model, the spreading dynamics were averaged over 10^4^ iterations of the SIR dynamics to derive each ESSES seizure pattern. The model fit analyses were repeated 10 times to obtain averaged values. For the Virtual resection analyses we performed 300 independent realizations of the seed regions and SIR dynamics. Each seed realization was used to measure seizure propagation in the original (before any resections) network and after the selected resection of each size. For the Optimization of resections analysis we also ran the simulated annealing algorithm 10 times for each resection size and selected the iteration that led to the minimal seed efficiency.

For the classification analyses we report the accuracy= (*T P* + *T N*)*/*(*T P* + *FP* + *FN* + *T N*), precision = *T P/*(*T P* + *FP*), sensitivity= *T P/*(*T P* + *FN*), *F* 1 statistic (harmonic mean between precision and sensitivity) = 2*T P/*(2*T P* + *FP* + *FN*), and area under the curve *AUC*. For the prediction analyses, we built a predictive model for each patient using the data from the remaining patients, in a leave-one-out crossvalidation-type setting. The predictive model compounded the prediction results from these *N* = 34 models. We measured its accuracy, precision, sensitivity and *F* 1 statistic.

Finally, for the Machine Learning analyses we used the Adaboost1 algorithm (Matlab 2018) using leave-one-out-cross-validation and default hyperparameters. The numbers of learners in each model was equal to the group size minus one (33 for the validation cohort and 48 for the combined cohort), and results were averaged over 10 iterations of the Adaboost1 algorithm for each classification model.

### 5.9 Data availability

The data used for this manuscript are not publicly available because the patients did not consent for the sharing of their clinically obtained data. Requests to access to the data-sets should be directed to the corresponding author. All user-developed codes are publicly available on Github https://github.com/anapmillan/computer_model_for_epilepsy.

## Supporting information

database_to_BNA

## Data Availability

The data used for this manuscript are not publicly available because the patients did not consent for the sharing of their clinically obtained data.
Requests to access to the data-sets should be directed to the corresponding author.

## 6 Acknowledgements

Ana P. Millán and Ida A. Nissen were supported by ZonMw and the Dutch Epilepsy Foundation, project number 95105006. Piet Van Mieghem has been funded by the European Research Council (ERC) under the European Union’s Horizon 2020 research and innovation programme (grant agreement No 101019718). The funding sources had no role in study design, data collection and analysis, interpretation of results, decision to publish, or preparation of the manuscript.

## Competing Interests

The authors declare that they have no competing interests.

## Author Contributions

A.P.M., E.C.W.S., C.J.S., I.A.N, A.H. conceptualized the study, E.C.W.S., C.J.S.,

I.A.N, S.I., J.C.B., P.V.M., A.H. participated in the funding acquisition, A.P.M, E.C.W.S., C.J.S, A.H. devised the Methodology, A.P.M. performed the formal analysis, A.P.M, I.A.N, C.J.S, A.H. devised the software and visualization E.C.W.S., C.J.S., P.V.M., A.H. participated in the supervision, E.C.W.S., S.I., J.C.B. provided resources, A.P.M. wrote the original draft and all authors participated in writing, review and editing.

## Notes

### Competing Interest Statement

The authors have declared no competing interest.

### Funding Statement

Ana P. Millan and Ida A. Nissen were supported by ZonMw and the Dutch Epilepsy Foundation, project number 95105006.
Piet Van Mieghem has been funded by the European
Research Council (ERC) under the European Union Horizon 2020 research and innovation programme (grant agreement No 101019718).

### Author Declarations

All patients gave written informed consent and the study was performed in accordance with the Declaration of Helsinki and approved by the VUmC (Vrij Universiteit Medical Center) Medical Ethics Committee.

