## Supplementary Information for "Individualized epidemic spreading models predict epilepsy surgery outcomes: a pseudo-prospective study"

[1]Amsterdam UMC location Vrije Universiteit Amsterdam, Department of Clinical Neurophysiology and MEG Center, De Boelelaan 1117, Amsterdam, The Netherlands
[2]Amsterdam UMC, Vrije Universiteit Amsterdam, Department of Neurosurgery, De Boelelaan 1117, Amsterdam, The Netherlands
[3]Amsterdam Neuroscience, Brain Imaging, Amsterdam, The Netherlands
[4]Amsterdam Neuroscience, Systems & Network Neurosciences, Amsterdam, The Netherlands
[5]Amsterdam Neuroscience, Neurodegeneration, Amsterdam, The Netherlands
[6]Amsterdam Neuroscience, Cancer Biology and Immonology, Amsterdam, The Netherlands
[7]Amsterdam Neuroscience, Imaging and Biomarkers, Amsterdam, The Netherlands
[8]Faculty of Electrical Engineering, Mathematics and Computer Science, Delft University of Technology, Delft, The Netherlands

#### 1 Patient data

Basic statistics of the patient groups are summarized in supplementary tables 1 (modeling cohort) and 2 (validation cohort).

#### 2 Individualized MEG brain networks

Seizure propagation was modeled on the patient-specific brain networks, as derived from MEG data, for both cohorts. One to three eyes-closed resting-state (supine position) MEG recordings of 10 to 15 minutes each were acquired for each patient in a magnetically shielded room (Vacuumschmelze GmbH, Hanau, Germany) during routine presurgical clinical practice (whole-head MEG system (Elekta Neuromag Oy, Helsinki, Finland) with 306 channels consisting of 102 magnetometers and 204 gradiometers). The data were sampled at 1250 Hz, and online filtered with an anti-aliasing filter at 410 Hz and a high-pass filter of 0.1 Hz. The head's position relative to the MEG sensors was determined using the signals from 4 to 5 head-localization coils that were recorded continuously. The positions of the head-localization coils and the outline of the scalp (roughly 500 points) were measured with a 3D digitizer (Fastrak, Polhemus, Colchester, VT, USA). The temporal extension of Signal Space Separation (tSSS) [1, 2] was used to remove artifacts using Maxfilter software (Elekta Neuromag, Oy; version 2.1), and the MEG data were filtered in the broadband (0.5 – 48.0 Hz). For a detailed description and parameter settings see Hillebrand et al. [3].

Pre-operative MRI scans (8 channel phased-array head coil on a 3T whole-body MR scanner, Discovery MR750, GE Healthcare, Milwaukee, Wisconsin, USA) were used for co-registration with the MEG data. Anatomical 3D T1-weighted images were obtained with a fast spoiled gradient-recalled echo sequence, and were interpolated to 1 mm isotropic resolution during reconstruction. The points on the scalp surface were used for co-registration of the MEG scans with the anatomical MRI of the patient through surface-matching. A single sphere was fitted to the outline of the scalp and used as a volume conductor model for the beamforming approach.

Neuronal activity was reconstructed using an atlas-based beamforming approach, modified from Hillebrand et al. [4], to reconstruct the time-series of neuronal activation of the ROI centroids [5]. We considered the 246 ROIs of the Brainetome (BNA) atlas [6], the centroids of which were inversely transformed to the co-registered MRI of the

| Case | Sex | Resection Area | $S_{\mathrm{RA}}$ | Engel Score | #E | #ECP | $N_{SR}$ |
|---|---|---|---|---|---|---|---|
| P1 | F | RF | 4 | 1A | 13 | 128 | 47 |
| P2 | F | RT, RO | 13 | 1A | 14 | 142 | 50 |
| P3 | F | LT, LO | 5 | 1A | 15 | 144 | 53 |
| P4 | M | RT | 13 | 1A | 13 | 126 | 49 |
| P5 | F | RT | 10 | 1A | 11 | 109 | 42 |
| **P6** | **F** | **RT** | **5** | **2A** | **9** | **99** | **40** |
| P7 | F | LT | 5 | 1A | 11 | 110 | 44 |
| P8 | F | LP | 4 | 1A | 10 | 104 | 37 |
| P9 | M | RT, RI, RP | 3 | 1A | 12 | 102 | 38 |
| **P10** | **F** | **RT** | **13** | **2D** | **11** | **114** | **45** |
| **P11** | **F** | **LF** | **4** | **2C** | **13** | **117** | **47** |
| P12 | M | LF | 6 | 1A | 12 | 124 | 40 |
| P13 | M | LT | 5 | 1A | 12 | 106 | 30 |
| **P14** | **F** | **LT** | **6** | **3A** | **15** | **194** | **60** |
| P15 | M | RT | 12 | 1A | 10 | 107 | 32 |

Supp. Table 1: Patient data (modellng cohort). Ep. = Epilepsy, y = years, $S_{\mathrm{RA}}$ = number of resected ROIs, #E = number of intracranial electrodes, #ECP = total number of electrode contact points, $N_{SR}$ = number of brain regions sampled by the SEEG electrodes. F = female (under "Sex"), M = male, R = right, L = left, F = Frontal lobe (under "Resection Area"), T = Temporal lobe, O = Occipital lobe, I = Insula, P = Parietal lobe.

patient. Then, a scalar beamformer (Elekta Neuromag Oy; beamformer; version 2.2.10) was applied to reconstruct each centroid's time-series, as detailed elsewhere [5]. The time-series of each centroid were visually inspected for epileptiform activity and artifacts. On average, 59.08 (range: $48-83$) 16384-sample interictal epochs of sufficient quality were selected for each patient. The epochs were further analyzed in Brainwave (version 0.9.151.5 [7]) and were down-sampled to 312 Hz, and filtered in the broadband (0.5 - 48 Hz).

### 3 iEEG Propagation Pattern

Patients in the modeling cohort underwent invasive EEG recordings using stereo-tactic electrode implantation as described also in [8]. The number and location of the intracerebral electrodes (Ad-Tech, Medical Instrument Corporation, USA, 10-15 contacts, 1.12 mm electrode diameter, 5 mm intercontact spacing; and DIXIE, 10-19 contacts, 0.8 mm electrode diameter, 2 mm contact length, 1.5 insulator length, $16-80.5$ insulator spacer length) were planned individually for each patient by the clinical team, and were based on the location of the hypothesized SOZ and seizure propagation pattern. The number of electrodes per patient varied between 9 and 15 (average = $12.1 \pm 1.8$) and the total number of contacts between 194 and 99 (average = $121 \pm 24$).

The locations of the electrode contact points were obtained for each patient from the post-implantation CT scan (containing the iEEG electrodes) that was co-registered to the preoperative MRI scan using FSL FLIRT (version 4.1.6) 12 parameter afine transformation. Each contact point was assigned the location of the nearest BNA ROI centroid. Given that BNA ROIs are in general larger than the separation between contact points, different contact points could be assigned to the same BNA ROI.

An iEEG seizure pattern was derived for each patient by a clinician (ECWvS). First, a representative seizure was chosen for each patient. Then, the onset time of ictal activity was identified for each iEEG channel, and the channels were sorted according to their onset times. In case that two channels became active at the same time, were assigned the same activation order. This activation pattern was then translated into the BNA space, so that the each sampled ROI $i$ was assigned an activation step. If channels with different onset times belonged to the same BNA ROI, the ROI was assigned the earliest of the possible times. This constituted the *iEEG seizure pattern.*

### 4 Seed-likelihood maps

In order to characterize the seed-likelihood maps, in Supp. figure 1 we show the fraction of seed-likelihood accounted for by RA nodes (panels A and E), the fraction of nodes with non-zero seed-likelihood that belonged to the RA (panels

| Case | Sex | Resection Area | $S_{RA}$ | Engel Score |
|---|---|---|---|---|
| 1 | F | RT | 5 | 1A |
| **2** | **M** | **LT** | **15** | **2A** |
| 3 | M | RT | 10 | 1A |
| 4 | F | LT | 22 | 1A |
| 5 | M | LT | 3 | 1A |
| **6** | M | **RT** | **11** | **2B** |
| 7 | M | RF | 2 | 1A |
| 8 | F | RT | 12 | 1A |
| **9** | **M** | **LF LP** | **3** | **4B** |
| 10 | F | RF | 4 | 1A |
| 11 | M | RT | 12 | 1A |
| 12 | M | RT | 11 | 1A |
| **13** | **M** | **LP** | **5** | **1B** |
| 14 | F | LT | 18 | 1A |
| 15 | M | RT | 10 | 1A |
| 16 | M | RT | 18 | 1A |
| 17 | F | LT | 19 | 1A |
| 18 | M | RT | 3 | 1A |
| 19 | M | LT | 4 | 1A |
| 20 | M | LT | 12 | 1A |
| **21** | **M** | **RF** | **10** | **3A** |
| 22 | F | LT | 19 | 1A |
| 23 | F | RT | 12 | 1A |
| 24 | F | LT | 17 | 1A |
| 25 | M | LT | 15 | 1A |
| **26** | **M** | **LT** | **13** | **2B** |
| 27 | M | LT | 10 | 1A |
| 28 | F | LT | 3 | 1A |
| 29 | F | LT | 10 | 1A |
| **30** | **M** | **RT** | **17** | **4A** |
| 31 | M | RT | 19 | 1A |
| 32 | F | LP | 2 | 1A |
| **33** | **F** | **LT** | **2** | **1B** |
| 34 | M | LT LP | 5 | 1A |

Supp. Table 2: Patient data (validation cohort). $S_{RA}$ = number of resected ROIs, F = female (under "Sex"), M = male, R = right, L = left, F = Frontal lobe (under "Resection Area"), T = Temporal lobe, O = Occipital lobe, I = Insula, P = Parietal lobe.

B and F) and the comparison of the RA seed-likelihood (panels C and G) and node (panels D and G) fractions between the SF and NSF groups, respectively for the modeling and validation cohort.

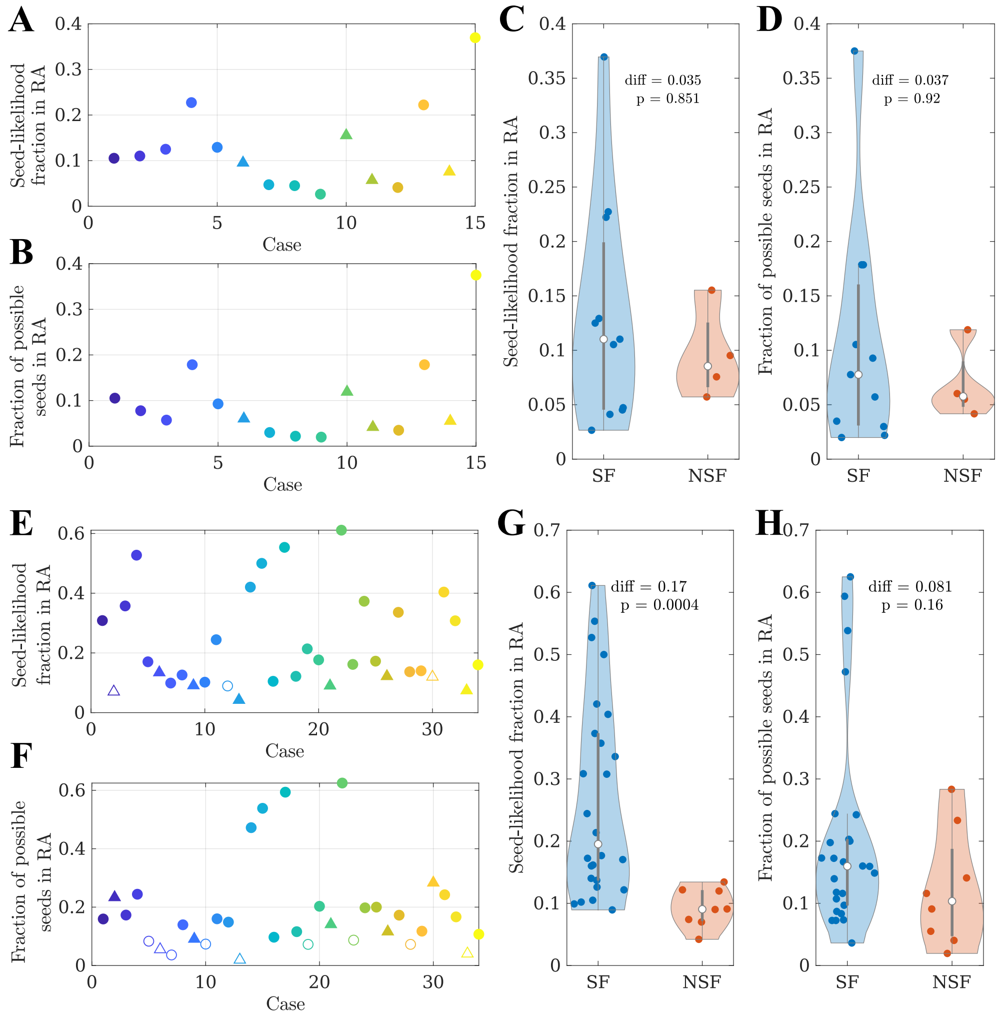


Supp. Figure 1: Characterization of seed-likelihood maps, modeling (top) and validation (bottom) cohorts. **A** and **E** Fraction of the total seed-likelihood in RA nodes, for each patient. Filled markers indicate that the seed-likelihood in the RA nodes was larger than expected by chance (this occurred for all patients in the modeling cohort). **B** and **F** Fraction of nodes with non-zero seed-likelihood that belonged to the RA. In **A** and **B**, SF (NSF) patients are indicated by circular (triangular) markers. The colorcode indicates the patient, also given by the x-axis. Filled markers indicate that there were more RA nodes with non-zero seed likelihood than expected by chance. **C,D** and **G,H** Comparison of the RA seed-likelihood (C) and node (D) fractions between the SF and NSF groups using a two-sided Wilcoxon ranksum test (panel C: diff=0.035, $p = 0.85$, $rks = 90$; panel D: diff=0.037, $p = 0.92$, $rks = 89$, panel G: diff=0.17, $p = 4 \cdot 10^{-5}$, $rks = 542$; panel H: diff=0.081, $p = 0.16$, $rks = 489.5$).

### 5 Seizure propagation model

#### 5.1 SIR Dynamics

The discrete-time SIR model [9, 10] was used to simulate the propagation of ictal activity from a set of *seed* nodes (or brain regions) that were set to be infected (or in the ictal state) at the beginning of the simulation. The SIR dynamics considered here are characterized by two parameters: the probability $\beta_{ij}$ that each infected node $i$ propagates the infection to a neighbour $j$ ($S \to I$) and the probability $\gamma_i$ that each infected node $i$ recovers ($I \to R$). For simplicity, and due to the limited amount of data available (one iEEG-recorded seizure per patient, and $N = 15$ patients) we considered here a global recovery rate $\gamma_i = \gamma$ and spreading probabilities given by the MEG network structure: $\beta_{ij} = w_{ij}$. As discussed in our previous studies [8, 11], the iEEG seizure patterns only accounted for the propagation of ictal activity and not the recovery to the healthy state, thus considering local recovery rates is beyond the scope of this study. In this manner we avoided introducing arbitrary time scales (i.e. to account for the duration of the ictal state) into the seizure propagation model.

The SIR dynamics was simulated by an adaptive Monte Carlo method (the BKL algoritihm) in Matlab in discrete time, such that at each time step one (and only one) new node became infected, and they were iterated $N_R = 10^4$ times per set of $(\rho, \gamma)$ parameters.

#### 5.2 Parameter fit: the ESSES model

The two control parameters of ESSES are the density of connections in the network, $\rho$, indirectly setting the spreading probabilities, and the global recovery rate $\gamma$. $\rho$ and $\gamma$ were fitted in the retrospective study (modeling cohort) by comparing the spatio-temporal propagation pattern of the ESSES-modeled seizures to the patient's iEEG seizure pattern (constructed as described above), when the resection area was used as the seed for epidemic spreading. For each set of ESSES model parameters, we measured the spatio-temporal seizure propagation profile described by the probability $p_i(t)$ that each ROI $i$ becomes infected at step $t$. Then, mean infection time $t_i$ of each ROI $i$ is defined as

$$t_i = \sum_{t=0}^{T} p_i(t), \tag{1}$$

where $T$ is the maximum integration time. $t_i$ was then sub-sampled to the ROIs sampled by the iEEG electrodes, and each of these ROIs was assigned an *infection order* according to $t_i$. This constituted the *ESSES seizure pattern.*

The *goodness-of-fit* $C(\rho, \gamma)$, quantified how similar the ESSES and iEEG seizure-propagation patterns were, and it was measured following the same procedure as in Ref. [8]. It took into account two factors: the weighted correlation between infection orders of ROIs that were infected in both patterns, $C_w$, and the overlap between the infected (or active) and susceptible (or inactive) ROI sets of both patterns, $P_{\text{overlap}}$, i.e.

$$C = C_w \cdot P_{\text{overlap}}. \tag{2}$$

In order to take into account the stochastic nature of the SIR dynamics, $C_w$ was weighted by the fraction of realizations that each ROI $i$ got infected during a modeled seizure, $P_{IR}(i)$. Similarly, $P_{\text{overlap}}$ was weighted by $P_{IR}(i)$ as:

$$P_{\text{overlap}} = N_{\text{iEEG}}^{-1} \left[ \sum_{i \in \mathbb{S}} P_{\text{IR}}(i) + \sum_{i \in \mathbb{H}} (1 - P_{\text{IR}}(i)) \right] = P_{\text{act}} + P_{\text{inact}}, \tag{3}$$

where $N_{\text{iEEG}}$ is the number of ROIs sampled by the iEEG electrodes (on average $= 43.6 \pm 7.9$), and $\mathbb{S}$ and $\mathbb{H}$ are respectively the sets of active (in the seizure state) and inactive (in the healthy state) ROIs in the iEEG pattern. The total correlation $C$ equals 1 in case of exactly equal activation patterns, 0 in the case of null-overlap or correlation, and $-1$ in the case of complete anti-correlation of activation times (but equal seizure areas). We note however that $C$ decays from 1 faster than a simple correlation metric, since it takes into consideration not only the activation times, but also the activation areas.

In order to fit $\rho$ and $\gamma$, we measured $C$ for a volume of the parameter space ($\gamma \in \left\{10^{-4}, 10^{-3}, 10^{-2}, 10^{-1}\right\}$, $\rho = \{0.025,\ 0.05,\ 0.10,\ 0.20,\ 0.30\}$) considering the resection area as the seed of seizure propagation [8, 11]. For each parameter configuration we performed 10 iterations (each over $N_R = 10^4$ iterations of the SIR dynamics), to the obtain average $C$ values and their fluctuation for each patient. We then found the parameter set that maximized $C$ for each patient (individual model fit) and at the group level (population model fit). The population model fit was used for all subsequent analyses in the main text (for both the modeling and validation cohorts). As in our previous study [8], we found that the population model fit led to a better classification between the SF and NSF groups, likely due to a reduced influence of noise.

The results for the individual model fit are summarized in figure 2 (on average, $C = 0.12$ with standard deviation $\text{std}_C = 0.07$). The model provided a (not significantly) better fit for the SF ($C_{SF} = 0.14 \pm 0.08$) than NSF ($C_{\text{NSF}} = 0.08 \pm 0.02$) patient groups ($C_{SF} - C_{\text{NSF}} = 0.05$, ranksum rks= 99, $p = 0.18$, exact two-sided Wilcoxon ranksum test), as shown in figure 2A. A ROC classification analysis based on the goodness-of-fit returned a good classification result with an area under the curve ($AUC$) of $AUC = 0.750$ (figure 2B). The confusion matrix corresponding to the optimal point of the ROC curve, according to the Youden criterion (to control for the class imbalance), is shown in 2C. We found an accuracy of 0.80, precision = 0.60, sensitivity = 0.75 and $F1 = 0.67$. There were no significant differences in the fit parameters $\rho$ and $\gamma$ between the groups.

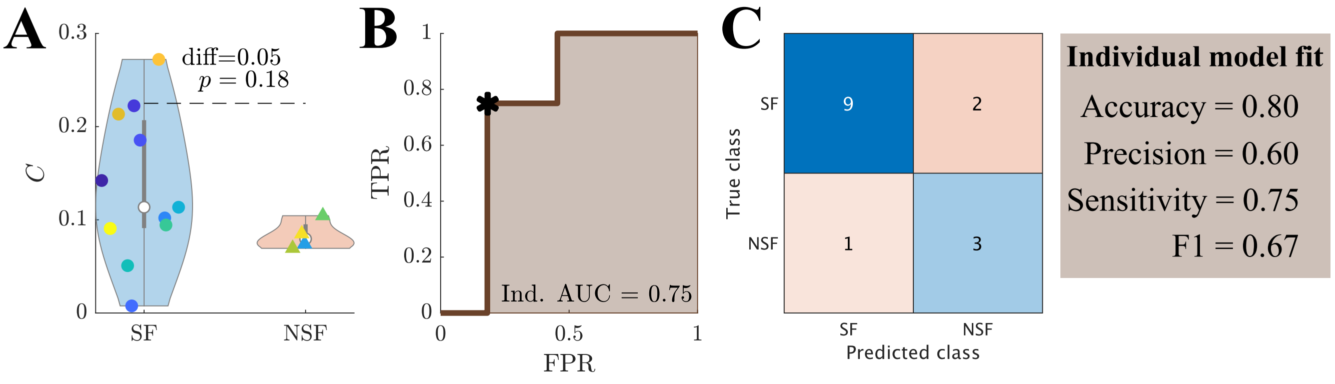


Supp. Figure 2: Seizure propagation model: model fit results. Group comparison of the goodness-of-fit using the individual model fit. **A** Violin plots of the goodness-of-fit distributions for each group. The significance analysis was performed with an exact two-sided Wilcoxon rank test. **B** ROC (receiving-operator-characteristic) curve analyses, where TPR and FPR respectively indicate the true positive (NSF patient classified as NSF) and false positive (SF patient classified as NSF) rates. **C** Confusion matrix corresponding to the optimal operating point of the ROC curve in panel B (shown by a black asterisk).

#### 5.3 Alternative resection strategies

We used an optimization method based on simulated annealing to find alternative resection strategies $R$ of increasing size $S(R)$. The optimization algorithm made use of the link between network structure and spreading dynamics to find resections that led to a minimum seizure propagation by minimizing the efficiency of the seed, $E_{\text{seed}}$. The seed in this case was defined as the set of regions with (significantly) non-zero seed-likelihood, and the contribution of each ROI was weighted by its seed-likelihood (see methods section for more details). At the population level, $E_{\text{seed}}(R)$ decreased with the size of the resection for all patients (see an exemplary case in figure **??**A-top, and the remaining cases in figure 4). To diminish differences due to seed extent and initial efficiency, we computed the normalized seed efficiency $e(\text{seed})_R = E(\text{seed})_0 - E(\text{seed})_R$ (figure 3B-top for the population average). At the group level, the SF group showed a significantly larger $e(\text{seed})_R$ than the NSF group ($F(19) = 14.80$, $p = 10^{-32}$), although the effect of increasing the resection size was not significantly different ($F(19) = 0.74$, $p = 0.8$).

The actual effect of each resection on seizure propagation in the model was quantified using the SIR dynamics and 300 independent seed realizations. An exemplary case is shown in supplementary figure 3A-bottom, and the group average can be seen in supplementary figure 3B-middle. Seizure propagation depended heavily on the seed realization. A bi-stable regime emerged where, depending on the seed realization, the modelled seizures either propagated macroscopically or died locally. For each resection size we measured the normalized decrease in seizure propagation $\delta IR(R)$. At the group level, the SF group had a significantly smaller $IR$ ($F(19) = 7.86$, $p = 10^{-16}$, supplementary figure 3-middle), but there was no significant difference in the effect of increasing the resection size ($F(19) = 0.29$, $p \approx 1$).

We defined the normalized decrease in seizure propagation to compare between different patients as $\delta IR(R) = (IR_0 - IR_R)/IR_0$ (figure 3B-bottom). At a group level, the SF group presented significantly larger $\delta IR(R)$ for

$S(R) > 1$ ($F(19) = 8.57$, $p = 2 \cdot 10^{-18}$), and again there was no significant difference in the effect of increasing the resection size between the groups ($F(19) = 0.27$, $p \approx 1$).

In order to compare the two groups systematically, we defined the optimal resection $R_{op}$ as the one leading to a 90% decrease in seizure propagation, $\delta IR = 0.90$, and the disconnecting resection $R_D$ as the smallest resection leading to seed disconnection, $e_{R_D} = 0$. We found that the SF group had smaller optimal resections (panel C-top) than the NSF group, but the difference was not significant (diff$= -5.73$, $rks = 81$ $p = 0.37$). The overlap between the optimal and actual resection was also larger, but not significantly so, for the SF group (panel D-top, diff$= 0.23$, $rks = 98$ $p = 0.20$). The disconnecting resection (panel E-top) was smaller, but not significantly so, for the SF than for the NSF group (diff$= -6.48$, $rks = 81$ $p = 0.37$). Either of these three variables could classify the two groups with good AUC results of 0.61, 0.66 and 0.61, respectively (mid subpanels of panels C, D and E). The confusion matrices corresponding to the optimal points (Youden criterion) are also shown in figure 3C,D,E. The classifications resulted in accuracy = 0.80, precision = 0.67, sensitivity = 0.50 and $F1 = 0.57$ for the size of optimal resection, accuracy = 0.67, precision = 0.44, sensitivity = 1.00 and $F1 = 0.62$ for the overlap of the optimal resections with the resection area, and accuracy = 0.80, precision = 0.67, sensitivity = 0.50 and $F1 = 0.57$ for the size of disconnecting resections.

##### 5.4 Disconnecting resection

The disconnecting resection $R_D$ was defined as the smallest resection leading to seed disconnection, i.e. to $E_R(\text{seed}) = 0$. It is computationally much faster to obtain than $R_{op}$ given that it does not require the simulation of the SIR spreading dynamics. As shown in Supp. figure 5E, the size of $R_D$ is almost identical (correlation coefficient = 0.99) to the size of $R_{op}$.

We also show in figure 5E the correlation among the 4 model biomarkers (size of optimal and disconnecting resections $S(R_{op})$ and $S(R_D)$, overlap between the optimal and actual resection $Ov(RA, R_{op})$ and (model-based) effect of the actual resection $\delta IR(RA)$. Only $S(R_{op})$ and $S(R_D)$ were strongly correlated.

#### 6 Simulation of the resection strategy: Modeling cohort

We made use of the population model and the seed-likelihood maps to simulate the effect of the clinical resection of each patient. The resection was simulated by disconnecting the resection area from the network, and the seed regions were derived probabilistically from the seed-likelihood maps. The effect of the resection was quantified as the relative decrease in seizure propagation due to the resection $\delta IR(RA) = (IR_0 - IR_R(RA))/IR_0$, where $IR_0$ and $IR_R(RA)$ stand respectively for the propagation before and after the resection (figure 6B). The seizure propagation and the effect of the resection depended strongly on the seed realization, so the same set of seeds were used before and after the resection to estimate $\delta IR(RA)$, and we considered 300 independent seed realizations to diminish noise effects.

The effect of the surgery was larger for the SF than the NSF group (figure 6C), although the group difference was not significant ($\delta IR(RA, SF) - \delta IR(RA, NSF) = 0.14$, $rks = 98$, $p = 0.22$). A ROC analysis using $\delta IR(RA)$ to classify the two groups found an AUC$= 0.73$ (center subpanel). The confusion matrix corresponding to the optimal classification point (Youden criterion, right subpanel) shows an accuracy of 0.73, precision = 0.50, sensitivity = 1.00 and $F1 = 0.67$. Again, in this case the classification analysis was able to identify all NSF patients correctly.

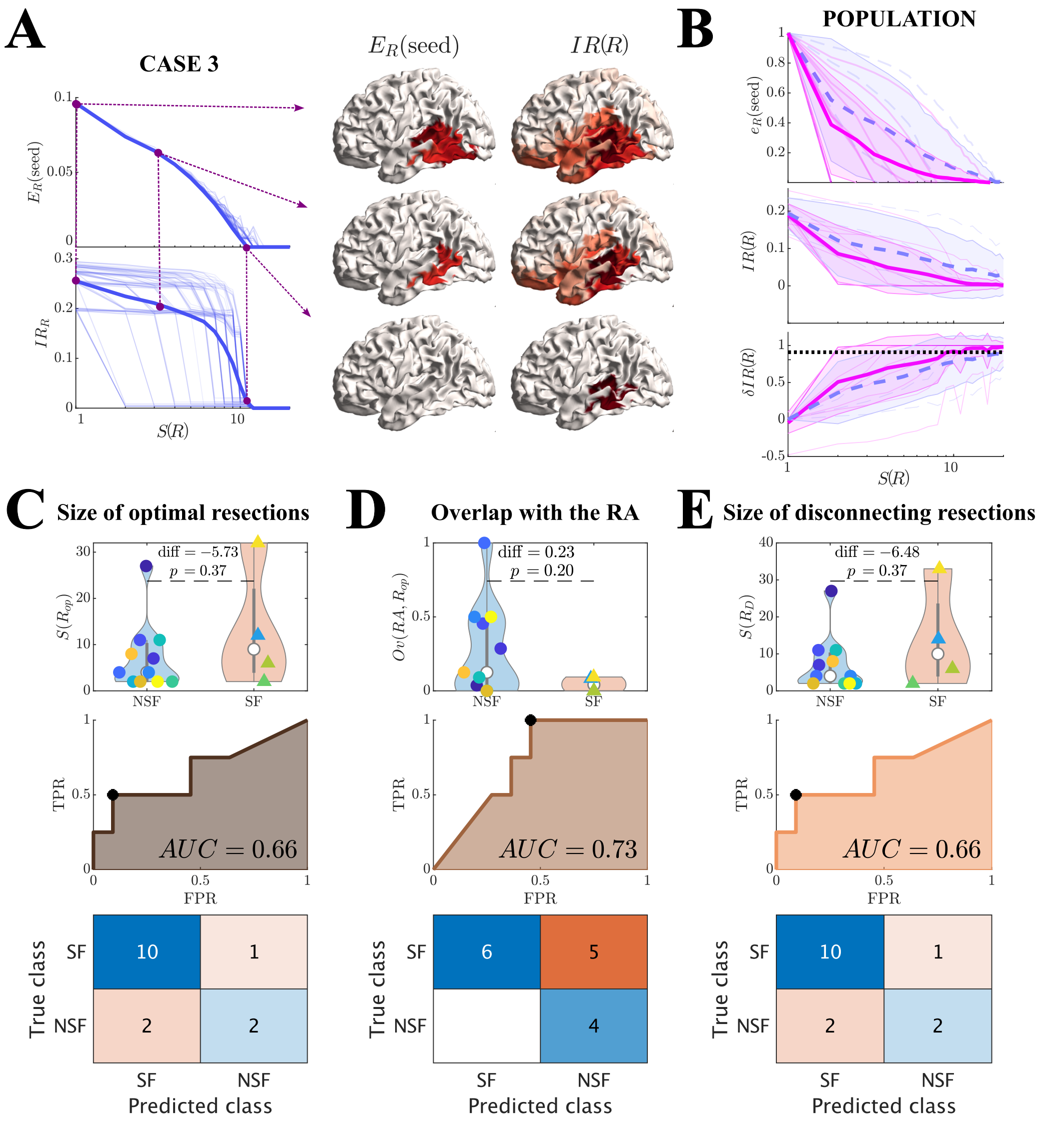


Supp. Figure 3: Optimization of virtual resections (modeling cohort). **A** Effect of resections of size $S(R)$ for an exemplary patient (case 3) as shown by the seed efficiency $E(\text{seed})_R$ as defined in the main text (top) and total seizure propagation $IR_R$ after the resection (bottom). Light lines in the top panel correspond to individual realizations of the optimization algorithm, and the solid wide line to the optimal resection of each size. This resection was used to quantify the effect on seizure propagation in the bottom panel. Light lines correspond each to a seed realization, whereas the solid wide line corresponds to the average over seed realizations. Brain plots on the right show $E(\text{seed})_R$ (left) and $IR_R$ (right) for three exemplary resection sizes ($S(R) = 1, 3, S_D$), where $S_D$ is the size of the disconnecting resection. **B** Group level analysis of the normalized seed efficiency $e(\text{seed})$, seizure propagation $IR_R$ and normalized decrease in seizure propagation $\delta IR(R)$ after (optimal) resections of size $S(R)$. Blue dashed lines and pink solid lines stand respectively for NSF and SF patients. Wide lines indicate the group averages with uncertainties shown by the shaded areas, as given by the standard deviation. The dotted black line in the bottom panel indicates the 90% threshold used to define the optimal resection. **C, D, E** Group level comparison of the size of optimal resections (C), its overlap with the actual resection area (D) and the size of disconnecting resections (E). The top panels illustrate the violin plots for the SF and NSF groups, with significance results given by exact two-sided Wilcoxon ranksum tests. Data-points are coded with the same colour and marker type as in supplementary figure 1A. The middle panels show the corresponding ROC classification analyses, where FPR and TPR stand respectively for the false positive and true positive rates. The bottom panels show the confusion matrices corresponding to the optimal classification points (Youden criterion).

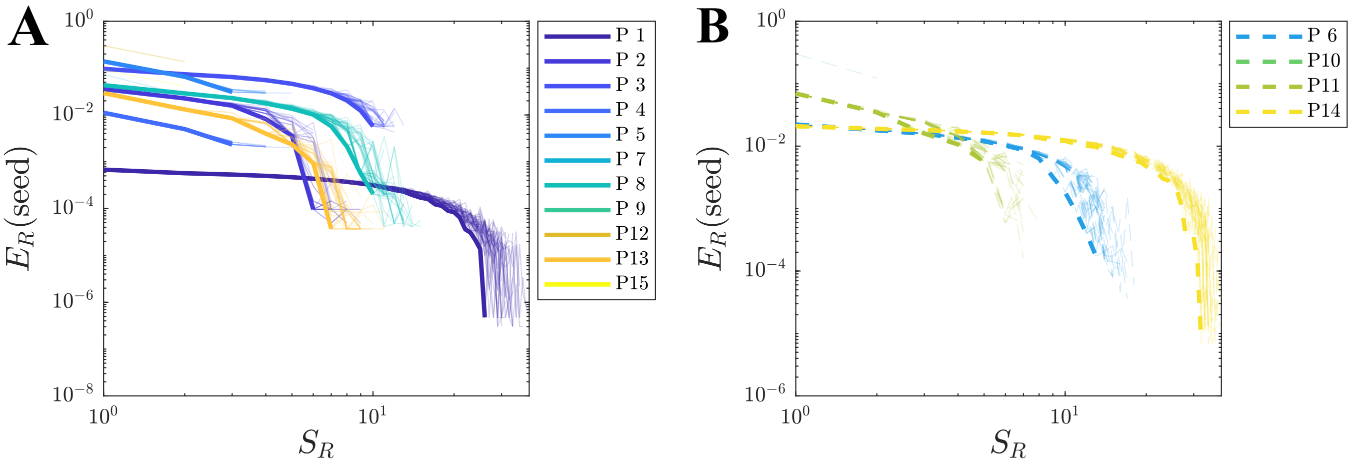


Supp. Figure 4: Optimization of virtual resection strategies for SF (panel A) and NSF (panel B) patients, showing the seed efficiency $E_R$ (seed) after virtual resections of size $S(R)$. The light lines indicate individual realizations of the simulated annealing algorithm, whereas the thick lines indicate the best iteration (i.e. the one leading to the minimum $E_R$ (seed). Each patient is shown by a different colour as indicated in the legends, with the same color code as figure 1A.

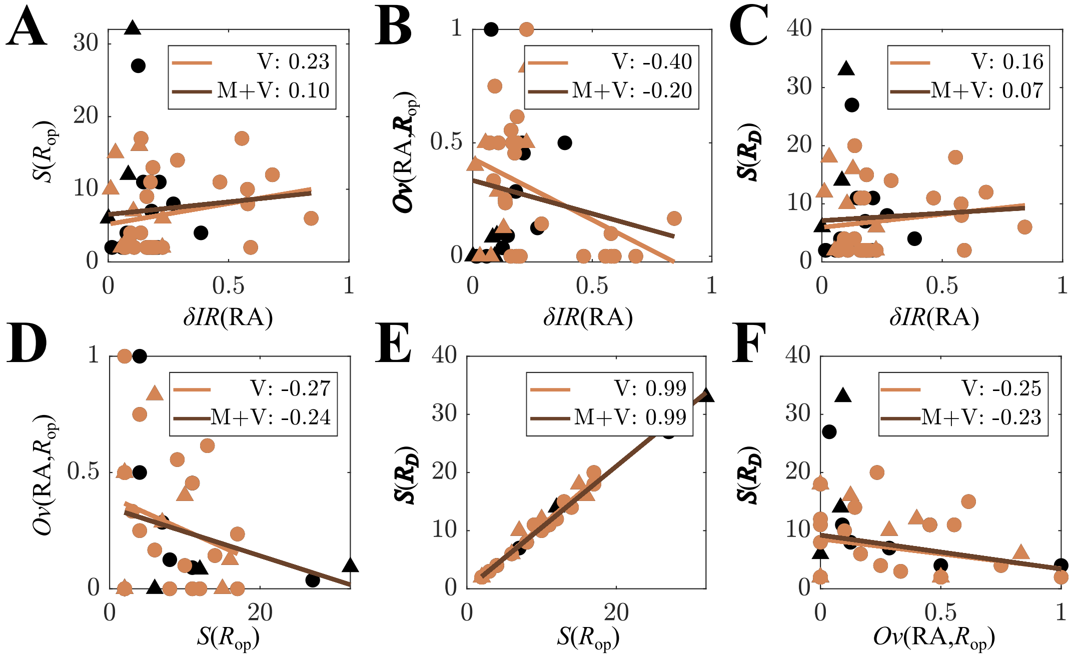


Supp. Figure 5: Pearson's correlation coefficient between each pair of model-derived variables: $\delta IR(RA)$, $S(R_{op})$, $Ov(RA, R_{op})$, $S(R_D)$. Black (dark) data-points stand for patients in the validation cohorts, whereas orange (light) markers stand for patients in the modelling cohort (thus the combined cohorts includes all data-points). The orange (light) lines indicate the linear correlation for the validation cohort, and the brown (dark) ones stand for the combined cohort $V + M$. The correlation coefficients are indicated in the legends.

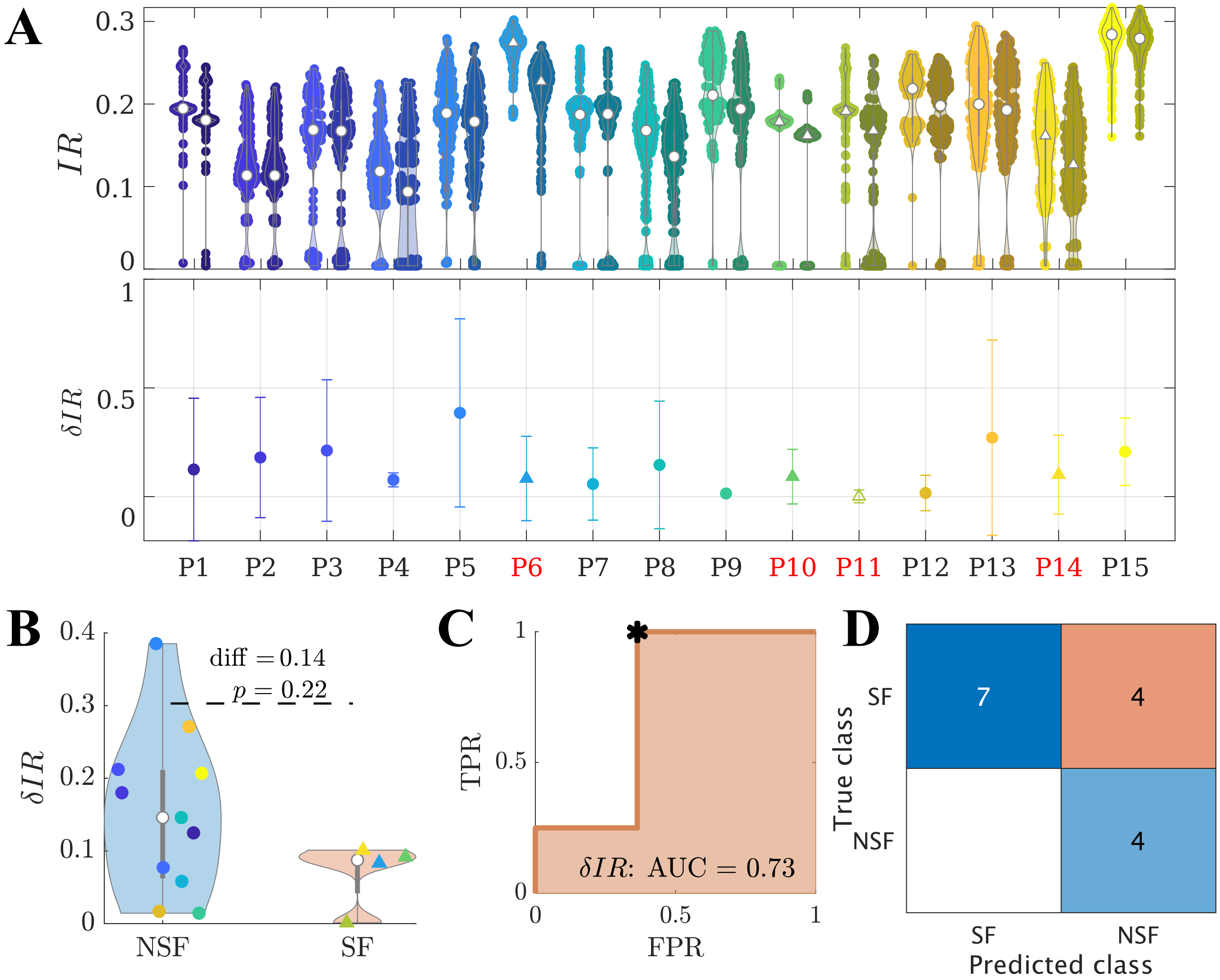


Supp. Figure 6: Simulation of the resective surgery (modelling cohort). **A** Effect of the virtual resections of the resection area in the model. The top panel shows the total seizure propagation $IR$ before and after the resection for each patient (respectively left and right violin plots for each x-tick), for 300 independent realizations of the seed regions. The white markers (respectively circles for SF and triangles for NSF cases) show the average over the seed realizations. NSF cases are also highlighted by red labels on the x-axis. The bottom panel shows the relative decrease in seizure propagation $\delta IR(RA)$ for each patient, as given by the average (data-points) and standard deviation (error bars). The colorcode is as in figure 1A. **B, C, D** Virtual resections of the resection area had a (not significantly) larger effect for the SF than NSF group, as shown by the violin plots of $\delta IR(RA)$ in panel B. A ROC classification analysis of the two groups based on $\delta IR(RA)$ showed an AUC$=0.73$ (panel C). TPR and FPR respectively stand for the true positive (NSF patients classified as NSF) and false positive (SF patients classified as NSF) rates. The confusion matrix corresponding to the optimal operating point (Youden criterion) is shown in panel D. For this threshold we found accuracy $= 0.73$, precision $= 0.50$, sensitivity $= 1.00$, and $F1 = 0.75$.
